# Collaborative Large Language Models for Automated Data Extraction in Living Systematic Reviews

**DOI:** 10.1101/2024.09.20.24314108

**Authors:** Muhammad Ali Khan, Umair Ayub, Syed Arsalan Ahmed Naqvi, Kaneez Zahra Rubab Khakwani, Zaryab bin Riaz Sipra, Ammad Raina, Sihan Zou, Huan He, Seyyed Amir Hossein, Bashar Hasan, R. Bryan Rumble, Danielle S. Bitterman, Jeremy L. Warner, Jia Zou, Amye J. Tevaarwerk, Konstantinos Leventakos, Kenneth L. Kehl, Jeanne M. Palmer, M. Hassan Murad, Chitta Baral, Irbaz bin Riaz

## Abstract

**Objective:** Data extraction from the published literature is the most laborious step in conducting living systematic reviews (LSRs). We aim to build a generalizable, automated data extraction workflow leveraging large language models (LLMs) that mimics the real-world two-reviewer process.

**Materials and Methods:** A dataset of 10 clinical trials (22 publications) from a published LSR was used, focusing on 23 variables related to trial, population, and outcomes data. The dataset was split into prompt development (n=5) and held-out test sets (n=17). GPT-4-turbo and Claude-3-Opus were used for data extraction. Responses from the two LLMs were compared for concordance. In instances with discordance, original responses from each LLM were provided to the other LLM for cross-critique. Evaluation metrics, including accuracy, were used to assess performance against the manually curated gold standard.

**Results:** In the prompt development set, 110 (96%) responses were concordant, achieving an accuracy of 0.99 against the gold standard. In the test set, 342 (87%) responses were concordant. The accuracy of the concordant responses was 0.94. The accuracy of the discordant responses was 0.41 for GPT-4-turbo and 0.50 for Claude-3-Opus. Of the 49 discordant responses, 25 (51%) became concordant after cross-critique, with an increase in accuracy to 0.76.

**Discussion:** Concordant responses by the LLMs are likely to be accurate. In instances of discordant responses, cross-critique can further increase the accuracy.

**Conclusion:** Large language models, when simulated in a collaborative, two-reviewer workflow, can extract data with reasonable performance, enabling truly ‘living’ systematic reviews.

## INTRODUCTION

As the medical field advances at an unprecedented rate, physicians struggle to keep up with the latest evidence to inform their practice.[1] Living systematic reviews (LSRs) address this challenge by continually integrating new findings and updating the evidence base.[2–9] However, the traditional LSR approach is susceptible to inefficiencies, with manual screening of citations and data extraction from publications being the most time-consuming tasks.[10] Additionally, error rates in manual data extraction can reach up to 50%, resulting from the diverse backgrounds and varying statistical and clinical expertise of data abstractors.[11] Moreover, manual data extraction is cost-intensive, adding to the challenge of maintaining LSRs in a truly ‘living’ state.[12] Automating the LSR workflow can enhance accuracy, reduce costs, and enable near-real-time evidence synthesis, addressing the critical bottleneck in maintaining LSRs’ timeliness.

Historically, natural language processing (NLP) techniques[13] such as Latent Dirichlet Allocation (LDA), Conditional Random Fields (CRFs), and support vector machines (SVM) were used for text mining unstructured publication text.[14–18] However, these models required extensively annotated datasets for training, consuming significant time and resources. The advent of transformer-based, pre-trained large language models (LLMs) has revolutionized text generation and processing with natural language instructions.[19] Large language models, capable of tasks such as named entity recognition (NER), relation extraction, event identification, and text summarization,[20] have been widely investigated for generalizable information extraction.[21] Pre-trained LLMs, enhanced through task-specific fine- and instruction-tuning,[22] show diverse capabilities in data extraction and other systematic review tasks without requiring large volumes of annotated datasets for training.[23–26] Notably, LLMs can critique and improve responses generated by other LLMs.[27] While commercial LLMs pretrained on the general domain can perform unsupervised data extraction without further fine-tuning, various model-, data- and framework-centric strategies can refine the response quality.[28] Among these strategies, prompt engineering has emerged as an effective technique to condition LLM responses[29] without the risk of overfitting associated with task-specific fine-tuning.[30]

Given the unmet need for efficient data extraction for LSRs, we investigate a generalizable workflow leveraging Open AI’s GPT-4-turbo (GPT-4T)[31] and Anthropic’s Claude-3-Opus (Claude-3O)[32] to automate data extraction from clinical trial publications. The workflow employs systematically designed prompts in a zero-shot experimental design, mimicking the traditional two-reviewer process in systematic reviews. We hypothesize that responses that are concordant between the LLMs are likely to be accurate, and in the instances of discordance, cross-critique between the LLMs will improve concordance.

## BACKGROUND AND SIGNIFICANCE

Utilizing LLMs for data extraction in systematic reviews and meta-analyses is an area of growing interest.[33] Kartchner et al. explored Open AI’s GPT-3.5-turbo and GPT-JT for summarizing and extracting information from clinical trials in two meta-analyses.[34] In a zero-shot setting, GPT-3.5-turbo outperformed GPT-JT, however, its use was limited by a high degree of hallucinations within the generated responses. Gartlehner et al. used Anthropic’s Claude-2 to extract data from 10 clinical trial publications, achieving an accuracy of 96.3%.[35] However, the study was limited to categorical variables and clinical trials with only one or two arms. Konet et al. compared GPT-4 and Claude-2 using the same publications and demonstrated a higher data extraction accuracy by GPT-4 (100%).[36] However, this study focused on a few data elements, limiting the generalizability of their approach. Reason et al. used GPT-4 for data extraction and code generation for network meta-analyses, achieving over 99% success by qualitative assessment.[37] However, their approach relied only on publication text for data extraction and extracted data for a few variables, limiting its broader applicability.

Although these efforts showcase the promise of LLMs to automate data extraction for systematic reviews, existing efforts fall short by focusing on extracting high-level information that is insufficient for analyses, relying only on text to extract information, and demonstrating suboptimal performance. Utilizing a single LLM may generate responses with hallucinations, raising concerns about accuracy. We, therefore, investigate the collaborative LLM approach mimicking the traditional two-reviewer LSR workflow for accurate data extraction.

## MATERIALS AND METHODS

### Data Sources

This study utilized data from a published LSR and network meta-analysis on first-line treatments for metastatic castration-sensitive prostate cancer (mCSPC).[3] A manually curated gold-standard dataset derived from the full-text PDFs of 22 original and follow-up publications across 10 clinical trials was used to benchmark the data extraction performance of LLMs (*Supplementary Table 1*). The dataset included 23 variables categorized as “trial characteristics,” “population characteristics,” and “meta-analysis outcomes” for both the overall population and subgroups based on metastasis burden and timing (*Supplementary Table 2*).

### Data Processing

The workflow of preprocessing clinical trial publications, data extraction, and post-processing is shown in **Figure 1**. Each PDF was processed using rule-based programming with conventional Python libraries. Text from the publications was extracted using PyMuPDF.[38] Non-essential sections, e.g., “references,” “acknowledgments,” “appendices,” “ORCID IDs,” “author contributions,” “affiliations,” and “declaration of interests” were programmatically removed to focus on the relevant content and reduce the size of input tokens. The preprocessed text was split into smaller chunks based on GPT-4T and Claude-3O token limits to accommodate publications with longer text. Tiktoken library[39] was used to manage token counts. To ensure continuity of context, a 1000-token overlap between chunks was maintained. Pages containing the tables and figures were identified by keywords (“table” and “figure”) and captured as images at 500 dots per inch (DPI) resolution.

**Figure 1:**
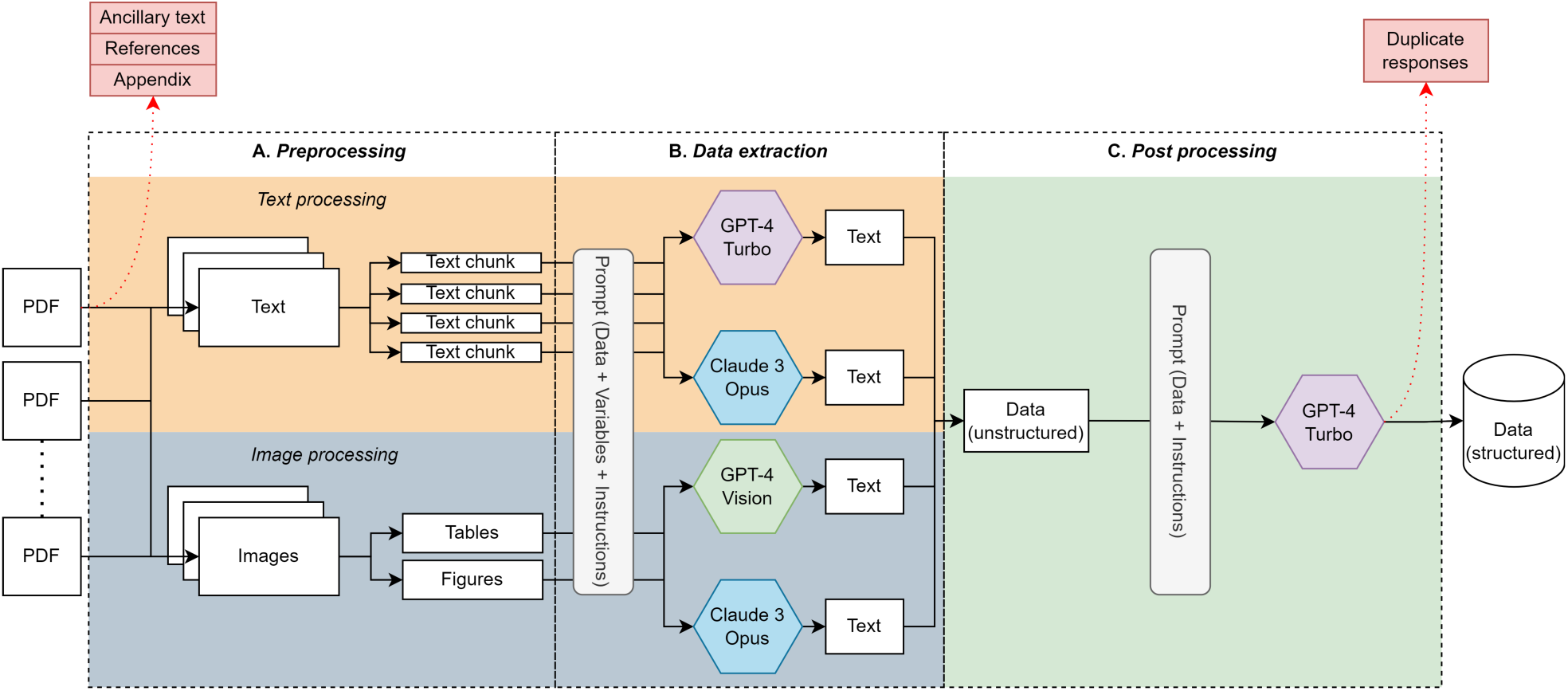
Data extraction workflow. Abbreviations: PDF: Portable document format The figure shows the data extraction approach used in our study. The approach includes (A). Preprocessing: Text is programmatically extracted from the PDF files of clinical trial publications, followed by the segregation into chunks. Ancillary text, including acknowledgments, references, and appendix, is removed. Tables and figures are identified based on the keyword-based search for “tables” and “figures” from the publications, with pages containing tables and figures captured as high-resolution images. (B). Data extraction: Extracted text chunks and images are provided to GPT-4-turbo and Claude-3-Opus, in a structured prompt comprising input text or image, variables, and instructions for data extraction. Responses generated by GPT-4-turbo and Claude-3-opus are in an unstructured format. (C). Post processing: Extracted data is converted in a structured format with GPT-4-turbo using a structured prompt that includes the unstructured data and formatting instructions. The duplicate responses from the structured responses are then removed.

### Sampling and Prompting

The dataset was randomly sampled at the publication level and split into prompt development and test sets. The prompt development set was used for iterative prompt engineering to extract relevant information from text and images (*Supplementary Figure 1*). The test set was held out and not evaluated until the prompts were finalized.

Chunks of text and images from each PDF were independently processed for data extraction using the application programming interfaces (APIs) of GPT-4T (*gpt-4-0125-preview* for text and *gpt-4-vision-preview* for images) and Claude-3O (*claude-3-opus-20240229*), the latest available versions as of April 2024. The temperature of the models was set to zero to generate deterministic responses. Initially, a baseline prompt was used, which was iteratively refined based on the prompt development set performance (*Supplementary Figure 1*). The final prompts included the input text or image, required variables, and user-defined instructions for data extraction (*Supplementary Methods*).

### Post-processing

Responses generated by GPT-4T and Claude-3O were processed separately. Extracted text and image data were concatenated and processed using GPT-4T in a structured format (*Supplementary Methods*). The duplicate responses from different text chunks and images of the same publication were removed. Likewise, the most frequent response was selected for a given variable in case of conflicting responses from different text chunks and images. The post-processing prompt included the extracted data and instructions for formatting.

### Concordance Assessment and Cross-critique

Responses from GPT-4T and Claude-3O were manually assessed for concordance. Numerical responses from GPT-4T and Claude-3O were required to match exactly to be considered concordant. Text-based responses from GPT-4T and Claude-3O were classified as concordant if they were identical or conveyed a similar meaning as defined by the expert reviewers. For discordant responses, the output of GPT-4T was provided to Claude-3O and vice versa for cross-critique. The cross-critique prompt included the original text or image, the other LLM’s response, and instructions to verify the response (*Supplementary Methods*). The responses after cross-critique were reassessed for concordance.

### Evaluation

Responses generated by GPT-4T and Claude-3O before and after concordance assessment and cross-critique were evaluated at the publication level for all variable categories. A sensitivity analysis was performed on the test set, excluding clinical trials that were also present in the prompt development set. LLM responses were manually compared to the gold standard, with performance metrics including accuracy, precision, recall, F1 score, and percentage of hallucinations calculated as mean values with 95% confidence intervals (CI). The Wilcoxon signed-rank test[40] was used to compare data extraction performance across different approaches, with a p-value of <0.05 indicating a statistically significant difference in performance.

### Data availability

The workflow for automated data extraction and the gold standard responses are available in a public repository (LLM extraction with cross-critique).

## RESULTS

Ten clinical trials with 22 publications were included in this study (*Supplementary Table 1*). The publications were randomly split, with 5 (23%) in the prompt development set and 17 (77%) in the test set. A total of 279,279 text tokens from the prompt development and test set publications were processed for data extraction, averaging 12,698 ± 3,972 tokens (range: 7,766 – 21,351) per publication. Additionally, 169 images were processed, averaging 7.6 ± 2 images (range: 3 – 12) per publication. The prompt development set had 65,999 text tokens (average: 13,199 ± 4,357; range: 9,502 – 18,975) and 42 images (average: 8 ± 2 images; range: 6 – 11). The test set had 213,370 text tokens (average: 12,551 ± 3,983; range: 7,766–21,351) and 127 images (average: 7 ± 2; range: 3–12).

A single LLM generated 506 responses for 23 variables across 22 publications (*Supplementary Figure 2*). The prompt development set had 115 responses, of which 110 (96%) were concordant and 5 (4%) were discordant. After cross-critique, 3 (60%) of the discordant responses became concordant, while 2 (40%) remained discordant. The test set had 391 responses, of which 342 (87%) were concordant and 49 (13%) were discordant. After cross-critique, 25 (51%) of the discordant responses became concordant, while 24 (49%) remained discordant.

### Response Evaluation in the Single LLM Approach

For 115 responses in the prompt development set, GPT-4T exhibited a mean accuracy of 0.95 (*95% CI: 0.93-0.97*), mean precision of 1.00, mean recall of 0.92 (*0.88-0.96*), and mean F1 score of 0.96 (*0.95-0.97*) whereas Claude-3O exhibited a mean accuracy of 0.95 (*0.93-0.97*), mean precision of 0.96 (*0.94-0.98*), mean recall of 0.97 (*0.95-0.99*) and mean F1 score of 0.96 (*0.94-0.98*). The evaluation of GPT-4T and Claude-3O by variable categories is reported in *Supplementary Table 3*. Hallucinations were present in none (0%) of the responses with GPT-4T and 2.60% (*95% CI: 0.00-6.08%*) of the responses with Claude-3O. The percentage hallucinations of GPT-4T and Claude-3O by variable categories are reported in *Supplementary Table 19*.

From 391 responses in the test set, GPT-4T exhibited a mean accuracy of 0.89 (*95% CI: 0.87-0.91*), mean precision of 0.98 (*0.96-1.00*), mean recall of 0.87 (*0.85-0.89*), and mean F1 score of 0.92 (*0.90-0.94*) whereas Claude-3O exhibited mean accuracy of 0.90 (*0.89-0.91*), mean precision of 0.94 (*0.92-0.96*), mean recall of 0.91 (*0.89-0.93*) and mean F1 score of 0.92 (*0.90-0.94*) (*Supplementary Table 3*). Hallucinations were present in 2.29% (*95% CI: 0.61-3.98%*) of the responses with GPT-4T and 2.76% (*0.27-5.26%*) of the responses with Claude-3O (*Supplementary Table 19*).

Sensitivity analyses showed GPT-4T to have a mean accuracy of 0.89 (*95% CI: 0.88-0.90*), mean precision of 0.98 (*0.97-0.99*), mean recall of 0.90 (*0.89-0.91*), and a mean F1 score of 0.93 (*0.92-0.94*) and Claude-3O to have a mean accuracy of 0.92 (*95% CI 0.91-0.93*), mean precision of 0.94 (*0.93-0.95*), mean recall of 0.95 (*0.94-0.96*), and a mean F1 score of 0.94 (*0.93-0.95*) (*Supplementary Table 9*). Hallucinations occurred in 2.00% (*95% CI 0.00%-4.59%*) of the responses with GPT-4T and 1.44% (*0.00%-3.49%*) of the responses with Claude-3O (*Supplementary Table 24*). The evaluation of responses by clinical trial publications is reported in *Supplementary Table 14*.

### Response Evaluation in the Collaborative LLM Approach

#### Concordant responses

From 115 responses in the prompt development set, GPT-4T and Claude-3O generated 110 (96%) concordant responses. The concordant responses exhibited a mean accuracy of 0.99 (*95% CI: 0.98-1.00*), mean precision of 1.00, mean recall of 0.96 (*0.94-0.97*), and mean F1 score of 0.98 (*0.97-0.99*). The evaluation of concordant responses by variable categories is reported in *Supplementary Table 4*. Hallucinations were present in none (0%) of the responses. The percentage hallucinations of concordant responses by variable categories are reported in *Supplementary Table 20*.

From 391 responses in the test set, GPT-4T and Claude-3O generated 342 (87%) concordant responses. The concordant responses exhibited a mean accuracy of 0.94 (*95% CI: 0.93-0.95*), mean precision of 1.00, mean recall of 0.92 (*0.90-0.94*), and mean F1 score of 0.96 (*0.956-0.964*) (*Supplementary Table 4*). Concordant responses significantly outperform GPT-4T in terms of the mean accuracy (p-value: 0.003), precision (0.035), recall and F1 score (0.003), and Claude-3O in terms of the mean accuracy (p-value: 0.007), precision (0.022) and F1 score (0.004). The comparison of concordant responses with GPT-4T and Claude-3O by variable categories is reported in *Supplementary Table 5.* A comparison of the test set data extraction accuracy by GPT-4T and Claude-3O in the single LLM approach and concordant responses in the collaborative LLM approach is shown in **Figure 2**. Comparison of the test set data extraction precision, recall, and F1 scores by GPT-4T and Claude-3O in the single LLM approach and concordant responses in the collaborative LLM approach are shown in *Supplementary Figures 3*, *4*, and *5*, respectively. Hallucinations were present in 0.25% *(95% CI: 0.00-0.70%*) of the responses (*Supplementary Table 20*).

**Figure 2:**
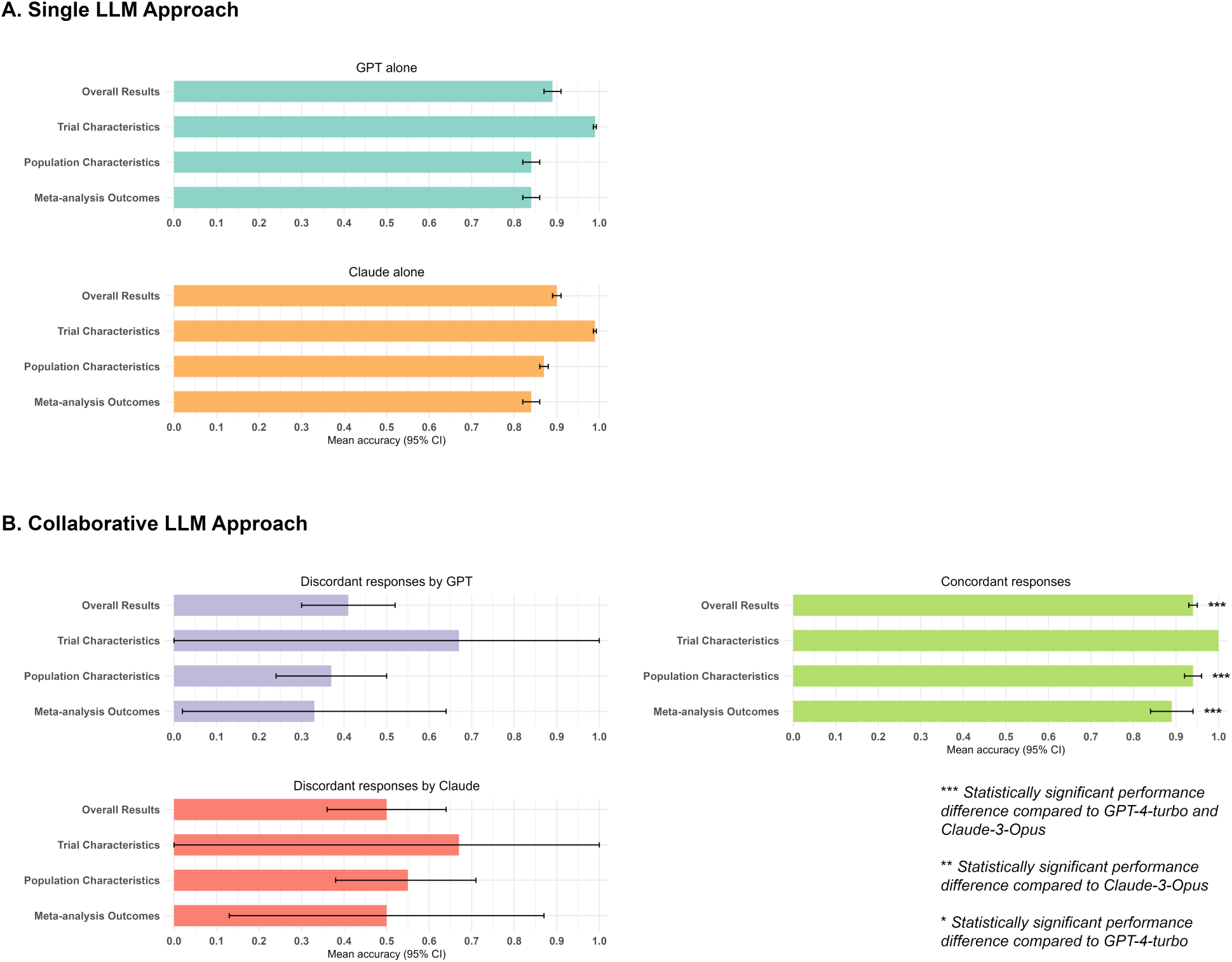
Mean accuracy of the individual versus the 2-reviewer collaborative LLM approach in the test set responses. Abbreviations: LLM: Large language model; GPT: Generative pretrained transformer The figure shows (A). Mean data extraction accuracy with 95% confidence intervals (CI) of GPT-4-turbo and Claude-3-Opus in the single LLM approach. (B). Mean data extraction accuracy with 95% CI of the discordant responses by GPT-4-turbo and Claude-3-Opus, and for concordant responses in the collaborative LLM approach. The concordant responses significantly outperform either GPT-4-turbo or Claude-3-Opus alone in overall data extraction accuracy and accuracy for extracting population characteristics and meta-analysis outcomes from clinical trial publications

Sensitivity analyses showed the concordant responses to have a mean accuracy of 0.97 (*95% CI: 0.96-0.98*), mean precision of 1.00, mean recall of 0.97 (*0.96-0.98*), and a mean F1 score of 0.98 (*0.975-0.984*) (*Supplementary Table 10*). Hallucinations occurred in none (0%) of the concordant responses (*Supplementary Table 25*). The evaluation of concordant responses by clinical trial publications is reported in *Supplementary Table 15*.

#### Discordant responses

From 115 responses in the prompt development set, GPT-4T and Claude-3O generated 5 (4%) discordant responses. In the subset of discordant responses, GPT-4T exhibited a mean accuracy of 0.25 (*95% CI: 0.00-0.69*), mean precision of 1.00, mean recall of 0.25 (*0.00-0.69*), and mean F1 score of 0.40 (*0.36-0.44*) whereas Claude-3O exhibited a mean accuracy of 0.50 (*0.06-0.93*), mean precision of 0.50 (*0.06-0.93*), mean recall of 1.00 and mean F1 score of 0.67 (*0.63-0.71*). The evaluation of discordant responses by GPT-4T and Claude-3O by variable categories is reported in *Supplementary Table 6*. Hallucinations were present in none (0%) of the responses with GPT-4T and 50% (95% CI: 0.00-100.00%) of the responses with Claude-3O. The percentage hallucinations of GPT-4T and Claude-3O-generated discordant responses by variable categories are reported in *Supplementary Table 21*.

From 391 responses in the test set, GPT-4T and Claude-3O generated 49 (13%) discordant responses. In discordant responses, GPT-4T exhibited a mean accuracy of 0.41 (*95% CI: 0.30-0.52*), mean precision of 0.54 (*0.40-0.68*), mean recall of 0.46 (*0.32-0.60*), and mean F1 score of 0.50 (*0.36-0.64*), whereas Claude-3O exhibited a mean accuracy of 0.50 (*0.36-0.64*), mean precision of 0.60 (*0.46-0.74*), mean recall of 0.72 (*0.61-0.83*) and mean F1 score of 0.65 (*0.54-0.76*) (**Figure 2**; *Supplementary Table 6; Supplementary Figures 3, 4*, and *5*). Hallucinations were present in 26.93% (*95% CI: 6.38-47.48%*) of the responses with GPT-4T and 41.00% (*17.41-64.59%*) of the responses with Claude-3O (*Supplementary Table 21*).

Sensitivity analyses of the discordant responses showed GPT-4T to have a mean accuracy of 0.60 (*95% CI: 0.43-0.77*), mean precision of 0.57 (*0.37-0.77*), mean recall of 0.48 (*0.30-0.67*) and a mean F1 score of 0.51 (*0.32-0.69*) and Claude-3O to have a mean accuracy of 0.44 (*95% CI: 0.00-0.18*), mean precision of 0.44 (*0.26-0.63*), mean recall of 0.67 (*0.49-0.85*) and a mean F1 score of 0.53 (*0.34-0.72*) (*Supplementary Table 11*). Hallucinations occurred in 14.33% (*95% CI: 0.00%-36.24%*) of the discordant responses with GPT-4T and 50.00% (*6.17%-98.83%*) of the discordant responses with Claude-3O (*Supplementary Table 26*). The evaluation of responses by clinical trial publications is reported in *Supplementary Table 16*.

### Response Evaluation after Cross-critique

#### Concordant responses after cross-critique

From 5 discordant responses in the prompt development set, GPT-4T and Claude-3O generated 3 (60%) concordant responses upon cross-critique with a mean accuracy, precision, recall, and F1 score of 1.00. The evaluation of concordant responses after cross-critique by variable categories is reported in *Supplementary Table* 7. Hallucinations were present in none (0%) of the responses. The percentage hallucinations of concordant responses after cross-critique by variable categories are reported in *Supplementary Table 22*.

From 49 discordant responses in the test set, GPT-4T and Claude-3O generated 25 (51%) concordant responses upon cross-critique with a mean accuracy of 0.76 (*95% CI: 0.60-0.92*), mean precision of 0.72 (*0.52-0.92*), mean recall of 0.65 (*0.49-0.81*) and mean F1 score of 0.68 (*0.52-0.84*) (*Supplementary Table 7*). The overall test set concordance and accuracy of the concordant responses after cross-critique relative to the discordant responses by GPT-4T and Claude-3O before cross-critique is shown in **Figure 3**. The test set precision, recall, and F1 score of the concordant responses after cross-critique relative to the discordant responses by GPT-4T and Claude-3O before cross-critique is shown in *Supplementary Figures 6, 7*, and *8*, respectively. Hallucinations were present in 8.30% (*95% CI: 0.00-19.35%*) of the responses (*Supplementary Table 22*).

**Figure 3:**
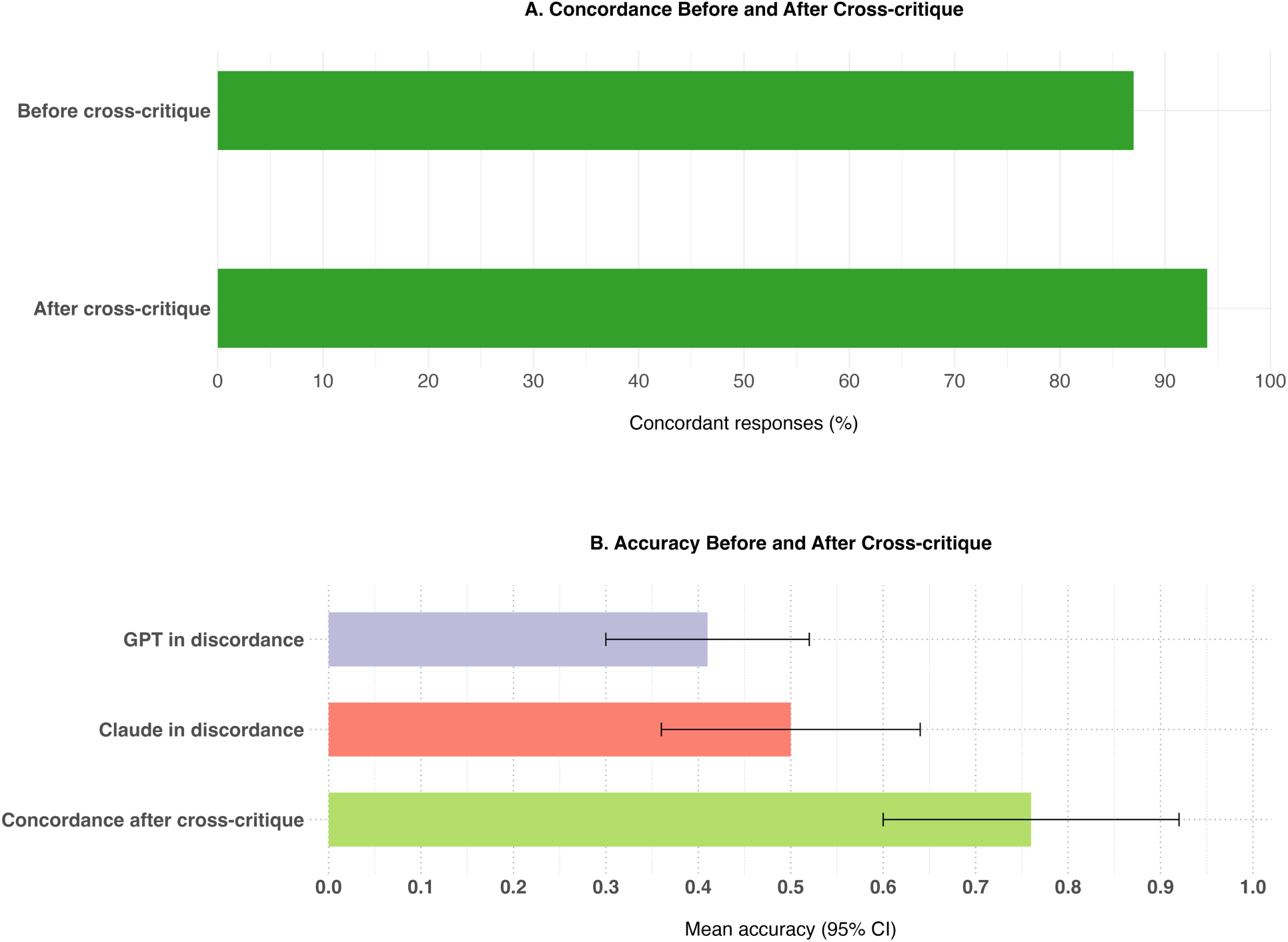
Cross-critique of discordant responses in the collaborative LLM approach. Abbreviations: GPT: Generative pre-trained transformer The figure shows (A). Overall percentage of test set concordant responses before and after cross-critique. Cross-critique of the discordant responses has shown to increase the overall test set concordance from 87% to 94% (B). Mean test set accuracy with 95% confidence intervals of the discordant responses by GPT-4-turbo and Claude-3-Opus, and the concordant responses after the cross-critique. Cross-critique has shown to generate concordant responses with a higher accuracy than the discordant responses by either GPT-4-turbo or Claude-3-Opus

Sensitivity analyses showed the concordant responses after cross-critique to have a mean accuracy of 0.85 (*95% CI: 0.75-0.95*), mean precision of 0.78 (*0.59-0.97*), mean recall of 0.63 (*0.45-0.81*) and a mean F1 score of 0.70 (*0.52-0.88*) (*Supplementary Table 12*). Hallucinations occurred in none (0%) of the concordant responses after cross-critique (*Supplementary Table 27*). The evaluation of concordant responses after cross-critique by clinical trial publications is reported in *Supplementary Table 17*.

#### Discordant responses after cross-critique

From 5 discordant responses in the prompt development set, GPT-4T and Claude-3O generated 2 (40%) discordant responses upon cross-critique. From the responses that were discordant after cross-critique, neither GPT-4T nor Claude-3O generated correct responses (mean accuracy: 0.00; mean precision: 0.00; mean recall: 0.00). The evaluation of discordant responses by GPT-4T and Claude-3O after cross-critique by clinical trial publications is reported in *Supplementary Table 8*. Hallucinations were present in 50% of the responses with GPT-4T and 100% of the responses with Claude-3O. The percentage hallucinations of GPT-4T and Claude-3O generated discordant responses after cross-critique by clinical trial reports is reported in *Supplementary Table 23*.

From 49 discordant responses in the test set, GPT-4T and Claude-3O generated 24 (49%) discordant responses upon cross-critique. From the responses that were discordant after cross-critique, GPT-4T exhibited a mean accuracy of 0.18 (*95% CI: 0.06-0.30*), mean precision of 0.29 (*0.13-0.45*), mean recall of 0.48 (*0.32-0.64*), and mean F1 score of 0.36 (*0.20-0.52*), whereas Claude-3O exhibited a mean accuracy of 0.45 (*95% CI: 0.29-0.61*), mean precision of 0.46 (*0.30-0.62*), mean recall of 0.75 (*0.59-0.91*) and mean F1 score of 0.57 (*0.41-0.73*) (*Supplementary Table 8*). Hallucinations were present in 56.25% (*95% CI: 30.57-81.93%*) of the responses with GPT-4T and 47.92% (*20.64-75.20%*) of the responses with Claude-3O (*Supplementary Table 23*).

Sensitivity analyses of the discordant responses after cross-critique showed GPT-4T to have a mean accuracy of 0.15 (*95% CI: 0.02-0.28*), mean precision of 0.38 (*0.10-0.65*), mean recall of 0.29 (*0.10-0.49*) and a mean F1 score of 0.33 (*0.10-0.56*) and Claude-3O to have a mean accuracy of 0.55 (*95% CI: 0.29-0.81*), mean precision of 0.42 (*0.14-0.69*), mean recall of 0.47 (*0.17-0.76*) and a mean F1 score of 0.44 (*0.15-0.73*) (*Supplementary Table 13*). Hallucinations occurred in 14.67% (*95% CI: 2.33%-81.00%*) of the discordant responses with GPT-4T and 50.00% (*6.17%-93.83%*) of the discordant responses with Claude-3O (*Supplementary Table 28*). The evaluation of responses by clinical trial publications is reported in *Supplementary Table 18*.

## DISCUSSION

This study simulated the real-world setting for data extraction in systematic reviews and showed that LLMs, when used collaboratively, analogous to the two-reviewer approach, demonstrate superior performance compared to the individual LLMs. The results of this study showed that the responses that are concordant between LLMs are likely to be correct. However, when they are discordant, cross-critique by LLMs, synonymous with human discussions while making decisions, further improves the accuracy. Given the rapid pace of evidence generation in domains like oncology[3–7] and COVID-19[41], the laborious data extraction process was the bottleneck for creating truly ‘living’ systematic reviews. Therefore, we have proposed a systematic LLM-based workflow (**Figure 4**) to simulate a two-reviewer setting that can be leveraged to facilitate the process of data extraction in living evidence synthesis informing clinical practice guidelines.

**Figure 4:**
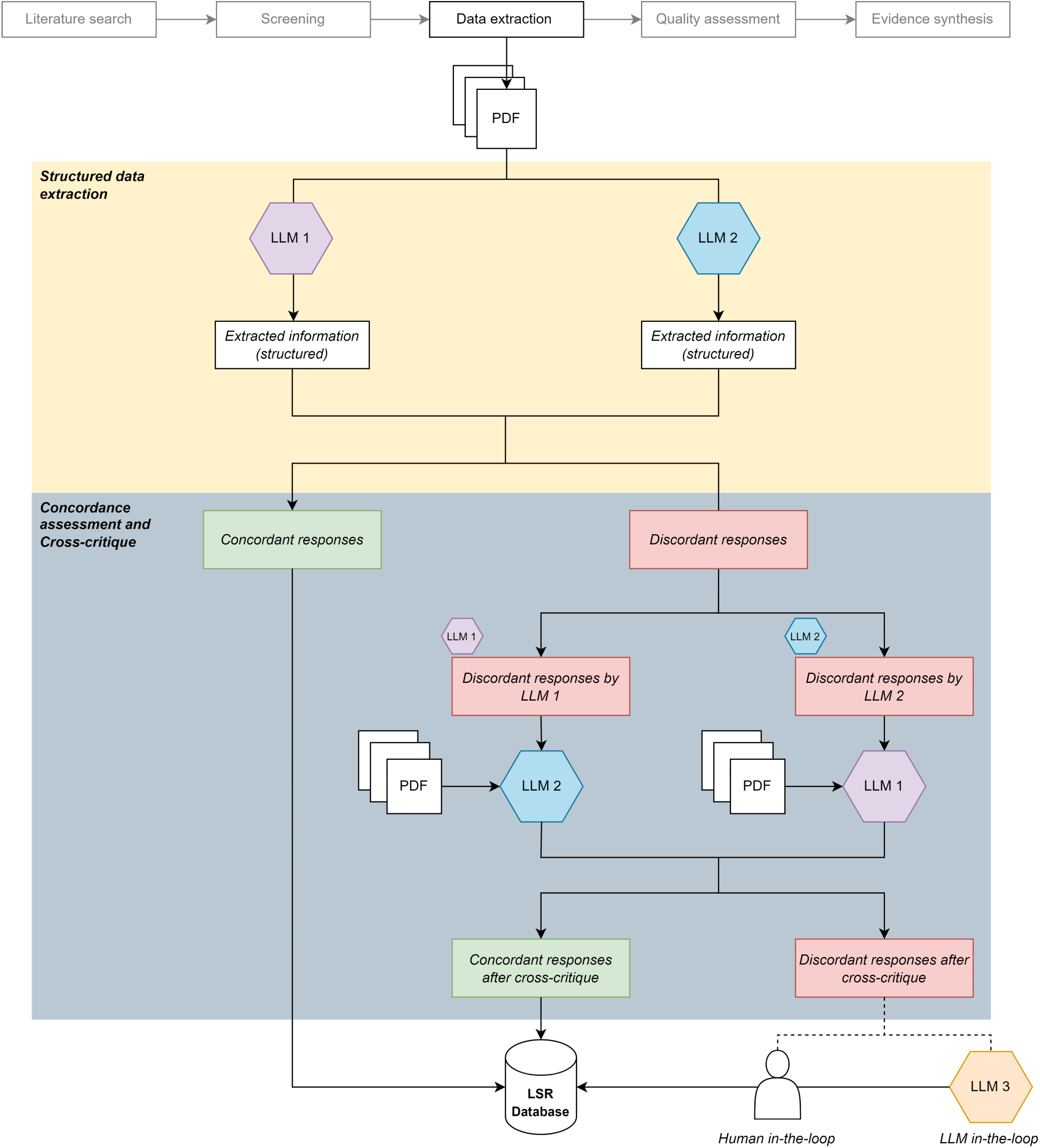
Implementation of the data extraction approach in the living evidence workflow. Abbreviations: PDF: portable document format; LLM: large language model; LSR: living systematic reviews The figure shows the implementation of the proposed collaborative LLM approach in the living evidence synthesis workflow. Analogous to the 2-reviewer manual approach, the 2-reviewer LLM is suggested for extracting data from clinical trial publications. Concordant from the collaborative LLM approach are selected as reliable, whereas the discordant responses are cross-critiqued. Concordant responses demonstrating a higher performance can be relied upon while the discordant responses require an additional human or an LLM-in-the-loop approach before being stored for downstream analyses.

The proposed approach has several strengths. First, the collaborative LLM approach reduces the risk of hallucinated responses, which is a major limitation of using individual LLMs in the real-world practice of systematic reviews. This is evident by a higher mean test set accuracy of 0.94 with collaborative LLMs compared to GPT-4T (0.89) and Claude-3O (0.90) alone (**Figure 2**). Likewise, the subset of responses with the collaborative LLM approach had fewer hallucinations (0.25%) compared to individual LLMs (∼2.5%). Second, we have developed a templated prompt specifically for data extraction that can be modified to extract different sets of variables across a variety of published articles (*Supplementary Figure 1*). This enables the reviewers to incorporate new variables in an existing LSR or to initiate a new review, making the workflow generalizable. Third, this approach offers significant time and cost savings. The average cost for data extraction was $3.40 for one publication, with an additional cost of $2.06 for cross-critique in almost real-time. Given these findings, this approach would significantly lower the costs and tremendously expedite the data extraction in LSR which otherwise would take months of manual effort[10 12].

However, the findings of this study should be interpreted in the context of some limitations. The dataset used in this study includes only prostate cancer trials and hence, the applicability to the fields outside oncology may be limited. However, we followed a systematic approach by randomly sampling our dataset into prompt development and test sets for unbiased evaluation. It could be argued that the inclusion of different publications from the same clinical trials in both prompt development and test sets could potentially contaminate the test data. However, a sensitivity analysis including unique clinical trials with no publications present in the prompt development set showed consistent results. It is also possible that the LLMs were previously exposed to publications that were included in the prompt development and test sets, which could have potentially overestimated the performance. While the collaborative approach achieved perfect accuracy for extracting variables like trial characteristics, the performance was relatively lower for other variables where implicit judgment is required, highlighting the need for more intelligent approaches for achieving automation. For example, introducing a third reviewer, either a human or an LLM, may mitigate this limitation.

In the future, we aim to integrate the proposed approach into our existing living interactive evidence (LIvE) synthesis framework[42] with a focus on reducing manual effort by iteratively identifying areas where LLMs can autonomously handle data synthesis tasks (**Figure 4**). Given the promising performance of the collaborative LLMs approach in this study, there is significant potential for its integration into both existing and new living evidence infrastructures.[3-7 43-54] Beyond its immediate application in data extraction, this approach could extend to other critical steps in the systematic review process, such as screening, quality assessment, and evaluating the certainty of evidence.[55] Additionally, the LLMs can be used collaboratively as planners for designing sophisticated analytic strategies enabling advanced meta-analysis.[56 57] Moreover, a key future direction involves further exploring the reasoning chains of LLMs, which could not only enhance confidence in their use but also improve trackability throughout the data extraction process. Beyond evidence synthesis, the collaborative LLM approach can be integrated into clinical workflows and systems to advance patient care and quality improvement initiatives. Developing guidelines and safeguards for using LLMs in evidence synthesis and healthcare is essential for widespread adoption. Training healthcare professionals and researchers to use AI-based tools effectively will be crucial for successfully implementing these solutions.

## CONCLUSIONS

In summary, concordant responses by the LLMs are likely to be accurate. In instances of discordant responses, cross-critique can further increase the accuracy. Large language models, when simulated in a collaborative, two-reviewer workflow, can extract data with reasonable performance, enabling truly ‘living’ systematic reviews. Validation of this workflow on external datasets is required to assess its widespread applicability.

## ACKNOWLEDGEMENTS

None

## COMPETING INTERESTS

Irbaz bin Riaz, Muhammad Ali Khan, Umair Ayub, Syed Arsalan Ahmed Naqvi, Kaneez Zahra Rubab Khakwani, Zaryab bin Riaz Sipra, Ammad Raina, Sihan Zou, Huan He, Seyyed Amir Hossein, Hasan Bashar, R. Bryan Rumble, Jia Zou, Kenneth L. Kehl, Jeanne Palmer, M. Hassan Murad and Chitta Baral do not have any relevant competing interests to disclose.

**Danielle S. Bitterman (DSB):** Editorial, unrelated to the submitted work: Associate Editor of Radiation Oncology, HemOnc.org (no financial compensation); Advisory and consulting, unrelated to the submitted work: MercurialAI

**Jeremy L. Warner (JLW):** Reports funding from AACR, NIH, Brown Physicians Incorporated, unrelated to the submitted work; consulting with Wested and The Lewin Group, unrelated to the submitted work; ownership in HemOnc.org LLC, unrelated to the submitted work

**Amye J. Tevaarwerk (AJT):** Family member at Epic Systems, unrelated to the submitted work

**Konstantinos Leventakos (KL):** Reports consulting activities (honoraria to institution) with Amgen, AstraZeneca Interdisciplinary Corporation, Boehringer Ingelheim Pharmaceuticals, Janssen Biotech, Novartis, unrelated to the submitted work; advisory boards (honoraria to institution) with AstraZeneca, Janssen, Jazz Pharmaceuticals, Mirati Therapeutics, Regeneron, Takeda, and Targeted Oncology, unrelated to the submitted work; CME activities (honoraria to institution) with OncLive and MJH Life Sciences, MD Outlook and Targeted Oncology, unrelated to the submitted work; Research support (to institution) from AstraZeneca and Mirati Therapeutics, unrelated to the submitted work.

## OTHER STATEMENTS

None

