## Supplementary File for "Collaborative Large Language Models for Automated Data Extraction in Living Systematic Reviews"

**Supplementary Material**

- Supplementary Methods
- Supplementary Table 1: Clinical trials assessed for data extraction
- Supplementary Table 2: Variable categories and corresponding variables for data extraction
- Supplementary Table 3: Mean evaluation metrics with 95% confidence intervals for responses across all publications
- Supplementary Table 4: Mean evaluation metrics with 95% confidence intervals for the subset of responses with concordance
- Supplementary Table 5: Wilcoxon signed-rank performance comparisons (p-value) of test set concordant responses with GPT-4-turbo and Claude-3-opus generated responses
- Supplementary Table 6: Mean evaluation metrics with 95% confidence intervals for the subset of responses with discordance
- Supplementary Table 7: Mean evaluation metrics with 95% confidence intervals for the subset of responses with concordance after cross-critique
- Supplementary Table 8: Mean evaluation metrics with 95% confidence intervals for the subset of responses with discordance after cross-critique
- Supplementary Table 9: Sensitivity analysis showing mean evaluation metrics with 95% confidence intervals for responses across all clinical trial publications included exclusively in the test set
- Supplementary Table 10: Sensitivity analysis showing mean evaluation metrics with 95% confidence intervals for responses with concordance across clinical trial publications included exclusively in the test set
- Supplementary Table 11: Sensitivity analysis showing mean evaluation metrics with 95% confidence intervals for responses with discordance across clinical trial publications included exclusively in the test set
- Supplementary Table 12: Sensitivity analysis showing mean evaluation metrics with 95% confidence intervals for responses with concordance after cross-critique across clinical trial publications included exclusively in the test set
- Supplementary Table 13: Sensitivity analysis showing mean evaluation metrics with 95% confidence intervals for responses with discordance after cross-critique across clinical trial publications included exclusively in the test set
- Supplementary Table 14: Mean evaluation metrics for responses by publications
- Supplementary Table 15: Mean evaluation metrics for a subset of responses with concordance by publications
- Supplementary Table 16: Mean evaluation metrics for a subset of responses with discordance by publications
- Supplementary Table 17: Mean evaluation metrics for a subset of responses with concordance after cross-critique by publications
- Supplementary Table 18: Mean evaluation metrics for a subset of responses with discordance after cross-critique by publications
- Supplementary Table 19: Mean percentage hallucinations with 95% confidence intervals for responses across all publications
- Supplementary Table 20: Mean percentage hallucinations with 95% confidence intervals for a subset of responses with concordance
- Supplementary Table 21: Mean percentage hallucinations with 95% confidence intervals for a subset of responses with discordance
- Supplementary Table 22: Mean percentage hallucinations with 95% confidence intervals for a subset of responses with concordance after cross-critique
- Supplementary Table 23: Mean percentage hallucinations with 95% confidence intervals for a subset of responses with discordance after cross-critique
- Supplementary Table 24: Sensitivity analysis showing mean percentage hallucinations with 95% confidence intervals for responses across all publications of clinical trials included exclusively in the test set
- Supplementary Table 25: Sensitivity analysis showing mean percentage hallucinations with 95% confidence intervals for responses with concordance across publications of clinical trials included exclusively in the test set
- Supplementary Table 26: Sensitivity analysis showing mean percentage hallucinations with 95% confidence intervals for responses with discordance across publications of clinical trials included exclusively in the test set
- Supplementary Table 27: Sensitivity analysis showing mean percentage hallucinations with 95% confidence intervals for responses with concordance after cross-critique across publications of clinical trials included exclusively in the test set
- Supplementary Table 28: Sensitivity analysis showing mean percentage hallucinations with 95% confidence intervals for responses with discordance after cross-critique across publications of clinical trials included exclusively in the test set
- Supplementary Figure 1: Prompt engineering workflow for data extraction
- Supplementary Figure 2: Breakdown of the generated responses
- Supplementary Figure 3: Mean precision of the individual versus the 2-reviewer collaborative LLM approach in the test set responses
- Supplementary Figure 4: Mean recall of the individual versus the 2-reviewer collaborative LLM approach in the test set responses
- Supplementary Figure 5: Mean F1 scores of the individual versus the 2-reviewer collaborative LLM approach in the test set responses
- Supplementary Figure 6: Mean precision of the test set discordant responses versus the concordant responses after cross-critique
- Supplementary Figure 7: Mean recall of the test set discordant responses versus the concordant responses after cross-critique
- Supplementary Figure 8: Mean F1 scores of the test set discordant responses versus the concordant responses after cross-critique

### **Supplementary Methods**

Prompt for data extraction:

A prompt for information extraction was designed through an iterative process of prompt engineering (*Supplementary Figure 1*). The prompt comprises of:

(i) A chunk of text or an image extracted from the PDF.
(ii) List of variables for which the information is required. The variables were defined in a spreadsheet, specifying the information to be extracted from reports. Although we used the sheet in .xlsx format, a .csv file can also be used. Variable information from the list was concatenated within the prompt. From a total of 23 variables, a subset of 5 variables was used in each prompt, and the responses were recorded. The number of variables within the subset was decided based on the prompt development set performance and can be altered based on the project requirements and the models’ performance for a particular systematic review. Since the required information can vary based on the requirements of a systematic review, the variables can be modified within the sheet.
(iii) Instructions specifying the data extraction format prompt the model to generate a concise response returning “NA” when the information is unavailable.

Prompt for post-processing:

A prompt for post-processing was engineered to arrange the extracted information in a structured format. The prompt comprised of:

(i) Response comprising extracted information.
(ii) Instructions to format the response as a colon (:) separating each variable from the information extracted for the corresponding variable.

Prompt for cross-critique:

A prompt for cross-critique was engineered to verify the response generated by GPT-4 Turbo (GPT-4T) by Claude-3 Opus (Claude-3O) and vice versa. The prompt comprised of:

(i) The chunk of text or image extracted from the PDF of the corresponding clinical trial report.
(ii) Information extracted for variables by another LLM.
(iii) Instructions to verify the response generated by the other LLM. If the verifying model identified the response as correct, it was instructed to state it. If the verifying model identified the response as incorrect, the model was prompted to generate the relevant response.

### **Supplementary Table 1: Clinical Trials Included in the Study**

| NCT | Name of the clinical trial | Number of publications | PMID(s) |
| --- | --- | --- | --- |
| NCT00268476 | STAMPEDE | 7 | 26719232; 28578639; 29529169; 31447077; 31560068; 35411939; 37142371 |
| NCT02446405 | ENZAMET | 2 | 31157964; 36990608 |
| NCT01715285 | LATITUDE | 2 | 28578607; 30987939 |
| NCT00309985 | CHAARTED | 2 | 26244877; 29384722 |
| NCT00104715 | GETUG-AFU 15 | 2 | 23306100; 26610858 |
| NCT01809691 | SWOG-1216 | 1 | 35446628 |
| NCT01957436 | PEACE-1 | 1 | 35405085 |
| NCT02489318 | TITAN | 2 | 31150574; 33914595 |
| NCT02677896 | ARCHES | 1 | 35420921 |
| NCT02799602 | ARASENS | 2 | 35179323; 36795843 |

Abbreviations: NCT: National clinical trial number; PMID: PubMed ID

The table shows national clinical trial numbers, names of the clinical trials, number of publications of each clinical trial and the PubMed IDs for all publication included in the study.

### **Supplementary Table 2: Variable categories and corresponding variables for data extraction**

| Variable Category | Variable |
| --- | --- |
| *Trial Characteristics* | Name of the clinical trial |
|  | Year of publication |
|  | Name of experimental arm |
|  | Name of control arm |
|  | Number of total participants included in the clinical trial |
|  | Median follow up duration in months of participants included in the clinical trial |
|  | Whether participants included in the clinical trial received docetaxel before or during the trial (Yes, No or NA) |
| *Population Characteristics* | Median age of participants included in experimental arm of the clinical trial |
|  | Median age of participants included in control arm of the clinical trial |
|  | Number of participants with high volume/burden metastasis included in the clinical trial |
|  | Number of participants with low volume/burden metastasis included in the clinical trial |
|  | Number of participants with synchronous metastasis (metastases at the time of diagnosis of prostate cancer) included in the clinical trial |
|  | Number of participants with metachronous metastasis (metastases after the diagnosis of prostate cancer) included in the clinical trial |
|  | Number of participants who have received the drug from treatment arm in the clinical trial |
|  | Number of participants with high volume/burden metastases (by CHAARTED criteria) who have received the drug from treatment arm in the clinical trial |
|  | Number of participants with low volume/burden metastases (by CHAARTED criteria) who have received the drug from treatment arm in the clinical trial |
|  | Number of participants with synchronous metastases (metastases at the time of diagnosis of prostate cancer) who have received the drug from treatment arm in the clinical trial |
|  | Number of participants with metachronous metastases (metastases after the diagnosis of prostate cancer) who have received the drug from treatment arm in the clinical trial |
| *Meta-analysis Outcomes* | Hazard ratio with confidence intervals of overall survival (OS) of all comparisons in all participants |
|  | Hazard ratio with confidence intervals of overall survival (OS) of all comparisons in all participants with high volume/burden metastases (by CHAARTED criteria) |
|  | Hazard ratio with confidence intervals of overall survival (OS) of all comparisons in all participants with low volume/burden metastases (by CHAARTED criteria) |
|  | Hazard ratio with confidence intervals of overall survival (OS) of all comparisons in all participants with synchronous metastases (metastases at the time of diagnosis of prostate cancer) |
|  | Hazard ratio with confidence intervals of overall survival (OS) of all comparisons in all participants with metachronous metastases (metastases after the diagnosis of prostate cancer) |

The table shows names of the variable categories with information regarding the corresponding variables. Data was extracted for all the variables in our study using a systematic prompt including information for the variables to be extracted. The variables were concatenated in the prompt, and the extracted information for each variable was recorded across clinical trial publications.

### **Supplementary Table 3: Mean evaluation metrics with 95% confidence intervals for responses across all publications**

**(a) GPT-4-turbo in a zero-shot setting**

| Variable Category | Accuracy | | Precision | | Recall | | F1 score | |
| --- | --- | --- | --- | --- | --- | --- | --- | --- |
|  | **Development** | **Test** | **Development** | **Test** | **Development** | **Test** | **Development** | **Test** |
| Overall Results | 0.95 (0.93-0.97) | 0.89 (0.87-0.91) | 1.00 (-) | 0.98 (0.96-1.00) | 0.92 (0.88-0.96) | 0.87 (0.85-0.89) | 0.96 (0.95-0.97) | 0.92 (0.90-0.94) |
| Trial Characteristics | 1.00 (-) | 0.99 (0.986-0.993) | 1.00 (-) | 1.00 (-) | 1.00 (-) | 0.99 (0.986-0.993) | 1.00 (-) | 0.99 (0.986-0.993) |
| Population Characteristics | 0.93 (0.91-0.95) | 0.84 (0.82-0.86) | 1.00 (-) | 0.95 (0.94-0.96) | 0.82 (0.78-0.86) | 0.83 (0.81-0.85) | 0.90 (0.89-0.91) | 0.89 (0.88-0.90) |
| Meta-analysis Outcomes | 0.92 (0.88-0.96) | 0.84 (0.82-0.86) | 1.00 (-) | 0.98 (0.97-0.99) | 0.92 (0.88-0.96) | 0.79 (0.76-0.82) | 0.96 (0.95-0.97) | 0.87 (0.85-0.89) |

**(b) Claude-3-opus in a zero-shot setting**

| Variable Category | Accuracy | | Precision | | Recall | | F1 score | | |
| --- | --- | --- | --- | --- | --- | --- | --- | --- | --- |
|  | **Development** | **Test** | **Development** | **Test** | **Development** | **Test** | | **Development** | **Test** |
| Overall Results | 0.95 (0.93-0.97) | 0.90 (0.89-0.91) | 0.96 (0.94-0.98) | 0.94 (0.92-0.96) | 0.97 (0.95-0.99) | 0.91 (0.89-0.93) | | 0.96 (0.94-0.98) | 0.92 (0.90-0.94) |
| Trial Characteristics | 1.00 (-) | 0.99 (0.986-0.993) | 1.00 (-) | 1.00 (-) | 1.00 (-) | 0.99 (0.986-0.993) | | 1.00 (-) | 0.99 (0.986-0.993) |
| Population Characteristics | 0.96 (0.95-0.97) | 0.87 (0.86-0.88) | 1.00 (-) | 0.91 (0.89-0.93) | 0.91 (0.88-0.94) | 0.88 (0.86-0.90) | | 0.95 (0.94-0.96) | 0.89 (0.87-0.91) |
| Meta-analysis Outcomes | 0.88 (0.84-0.92) | 0.84 (0.82-0.86) | 0.87 (0.83-0.91) | 0.91 (0.89-0.93) | 1.00 (0.0) | 0.86 (0.83-0.89) | | 0.93 (0.92-0.94) | 0.88 (0.86-0.90) |

Mean values with corresponding 95% confidence intervals for accuracy, precision, recall, and F1 score of (a) GPT-4-turbo and (b) Claude 3-opus for all the responses of all publications in a zero-shot setting

### **Supplementary Table 4: Mean evaluation metrics with 95% confidence intervals for the subset of responses with concordance**

| Variable Category | Responses* | | Accuracy | | Precision | | | Recall | | F1 score | |
| --- | --- | --- | --- | --- | --- | --- | --- | --- | --- | --- | --- |
|  | **Development** | **Test** | **Development** | **Test** | **Development** | **Test** | **Development** | | **Test** | **Development** | **Test** |
| Overall Results | 110 | 342 | 0.99 (0.98-1.00) | 0.94 (0.93-0.95) | 1.00 (-) | 1.00 (-) | 0.96 (0.94-0.97) | | 0.92 (0.90-0.94) | 0.98 (0.97-0.99) | 0.96 (0.956-0.964) |
| Trial Characteristics | 35 | 116 | 1.00 (-) | 1.00 (-) | 1.00 (-) | 1.00 (-) | 1.00 (-) | | 1.00 (-) | 1.00 (-) | 1.00 (-) |
| Population Characteristics | 53 | 151 | 0.96 (0.93-0.99) | 0.94 (0.92-0.96) | 1.00 (-) | 1.00 (-) | 0.88 (0.83-0.93) | | 0.92 (0.90-0.94) | 0.94 (0.93-0.95) | 0.96 (0.95-0.97) |
| Meta-analysis Outcomes | 22 | 75 | 1.00 (-) | 0.89 (0.84-0.94) | 1.00 (-) | 1.00 (-) | 1.00 (-) | | 0.84 (0.77-0.91) | 1.00 (-) | 0.91 (0.90-0.92) |

*Number of concordant responses generated for each variable category across all publications

Mean values with corresponding 95% confidence intervals for accuracy, precision, recall, and F1 score for the subset of responses where GPT-4-turbo and Claude-3-opus generated concordant responses

### **Supplementary Table 5: Wilcoxon signed-rank performance comparisons (p-value) of test set concordant responses with GPT-4-turbo and Claude-3-opus generated responses**

**(a) Concordant vs GPT-4-turbo**

| Variable Category | Accuracy | Precision | Recall | F1 score |
| --- | --- | --- | --- | --- |
| Overall Results | **0.003** | **0.035** | **0.021** | **0.003** |
| Trial Characteristics | 0.990 | - | 0.990 | 0.990 |
| Population Characteristics | **0.005** | **0.058** | **0.042** | **0.006** |
| Meta-analysis Outcomes | **0.005** | 0.058 | **0.042** | **0.006** |

**(b) Concordant vs Claude-3-opus**

| Variable Category | Accuracy | Precision | Recall | F1 score |
| --- | --- | --- | --- | --- |
| Overall Results | **0.007** | **0.022** | 0.527 | **0.004** |
| Trial Characteristics | 0.990 | - | 0.990 | 0.990 |
| Population Characteristics | **0.033** | 0.100 | 0.402 | 0.066 |
| Meta-analysis Outcomes | **0.033** | 0.100 | 0.402 | 0.066 |

p-values from Wilcoxon signed-rank tests comparing mean evaluation metrics of concordant responses with (a) GPT-4-turbo and (b) Claude-3-opus. Bold values indicate statistically significant improvements in concordant responses

### **Supplementary Table 6: Mean evaluation metrics with 95% confidence intervals for the subset of responses with discordance**

**(a) GPT-4-turbo in a zero-shot setting:**

| Variable Category | Responses* | | | Accuracy | | | Precision | | | Recall | | F1 score | |
| --- | --- | --- | --- | --- | --- | --- | --- | --- | --- | --- | --- | --- | --- |
|  | **Development** | **Test** | **Development** | | **Test** | **Development** | | **Test** | **Development** | | **Test** | **Development** | **Test** |
| Overall Results | 5 | 49 | 0.25 (0.00-0.69) | | 0.41 (0.30-0.52) | 1.00 (-) | | 0.54 (0.40-0.68) | 0.25 (0.00-0.69) | | 0.46 (0.32-0.60) | 0.40 (0.36-0.44) | 0.50 (0.36-0.64) |
| Trial Characteristics | 0 | 3 | - | | 0.67 (0.00-1.00) | - | | 1.00 (-) | - | | 0.67 (0.00-1.00) | - | 0.80 (0.75-0.85) |
| Population Characteristics | 2 | 36 | 0.00 (-) | | 0.37 (0.24-0.50) | - | | 0.55 (0.39-0.71) | 0.00 (-) | | 0.47 (0.31-0.63) | - | 0.51 (0.35-0.67) |
| Meta-analysis Outcomes | 3 | 10 | 0.50 (0.00-1.00) | | 0.33 (0.02-0.64) | 1.00 (-) | | 0.50 (0.06-0.93) | 0.50 (0.00-1.00) | | 0.17 (0.00-0.36) | 0.67 (0.62-0.72) | 0.25 (0.00-0.50) |

**(b) Claude-3-opus in a zero-shot setting**

| Variable Category | Responses* | | Accuracy | | Precision | | Recall | | F1 score | |
| --- | --- | --- | --- | --- | --- | --- | --- | --- | --- | --- |
|  | **Development** | **Test** | **Development** | **Test** | **Development** | **Test** | **Development** | **Test** | **Development** | **Test** |
| Overall Results | 5 | 49 | 0.50 (0.06-0.93) | 0.50 (0.36-0.64) | 0.50 (0.06-0.93) | 0.60 (0.46-0.74) | 1.00 (-) | 0.72 (0.61-0.83) | 0.67 (0.63-0.71) | 0.65 (0.54-0.76) |
| Trial Characteristics | 0 | 3 | - | 0.67 (0.00-1.00) | - | 1.00 (-) | - | 0.67 (0.00-1.00) | - | 0.80 (0.75-0.85) |
| Population Characteristics | 2 | 36 | 1.00 (-) | 0.55 (0.38-0.71) | 1.00 (-) | 0.68 (0.52-0.84) | 1.00 (-) | 0.71 (0.55-0.87) | 1.00 (-) | 0.69 (0.53-0.85) |
| Meta-analysis Outcomes | 3 | 10 | 0.00 (-) | 0.50 (0.13-0.87) | 0.00 (-) | 0.50 (0.13-0.87) | - | 1.00 (-) | - | 0.67 (0.65-0.69) |

*Number of discordant responses generated for each variable category across all publications

Mean values with corresponding 95% confidence intervals for accuracy, precision, recall, and F1 score of (a) GPT-4-turbo and (b) Claude-3-opus for the subset of responses where GPT-4-turbo and Claude-3-opus generated discordant responses

### **Supplementary Table 7: Mean evaluation metrics with 95% confidence intervals for the subset of responses with concordance after cross-critique**

| Variable Category | Responses* | | Accuracy | | Precision | | Recall | | | F1 score | |
| --- | --- | --- | --- | --- | --- | --- | --- | --- | --- | --- | --- |
|  | **Development** | **Test** | **Development** | **Test** | **Development** | **Test** | **Development** | **Test** | **Development** | | **Test** |
| Overall Results | 3 | 25 | 1.00 (-) | 0.76 (0.60-0.92) | 1.00 (-) | 0.72 (0.52-0.92) | 1.00 (-) | 0.65 (0.49-0.81) | 1.00 (-) | | 0.68 (0.52-0.84) |
| Trial Characteristics | 0 | 2 | - | 0.83 (0.41-1.00) | - | 1.00 (-) | - | 0.83 (0.41-1.00) | - | | 0.90 (0.84-0.96) |
| Population Characteristics | 2 | 20 | 1.00 (-) | 0.75 (0.57-0.93) | 1.00 (-) | 0.78 (0.60-0.96) | 1.00 (-) | 0.65 (0.43-0.87) | 1.00 (-) | | 0.71 (0.53-0.89) |
| Meta-analysis Outcomes | 1 | 3 | 1.00 (-) | 0.67 (0.00-1.00) | 1.00 (-) | 0.33 (0.00-1.00) | 1.00 (-) | 0.50 (0.00-1.00) | 1.00 (-) | | 0.40 (0.00-1.00) |

*Number of concordant responses generated for each variable category across all publications after cross-critique

Mean values with corresponding 95% confidence intervals for accuracy, precision, recall, and F1 score for the subset of responses where GPT-4-turbo and Claude-3-opus generated concordant responses after cross-critique

### **Supplementary Table 8: Mean evaluation metrics with 95% confidence intervals of the subset of responses with discordance after cross-critique**

**(a) GPT-4-turbo after cross-critique**

| Variable Category | Responses* | | Accuracy | | Precision | | Recall | | F1 score | |
| --- | --- | --- | --- | --- | --- | --- | --- | --- | --- | --- |
|  | **Development** | **Test** | **Development** | **Test** | **Development** | **Test** | **Development** | **Test** | **Development** | **Test** |
| Overall Results | 2 | 24 | 0.00 (-) | 0.18 (0.06-0.30) | 0.00 (-) | 0.29 (0.13-0.45) | 0.00 (-) | 0.48 (0.32-0.64) | - | 0.36 (0.20-0.52) |
| Trial Characteristics | 0 | 1 | - | 0.00 (-) | - | 0.00 (-) | - | - | - | - |
| Population Characteristics | 0 | 16 | - | 0.25 (0.05-0.45) | - | 0.46 (0.22-0.70) | - | 0.43 (0.23-0.63) | - | 0.44 (0.24-0.64) |
| Meta-analysis Outcomes | 2 | 7 | 0.00 (-) | 0.08 (0.00-0.23) | - | 0.08 (0.00-0.23) | 0.00 (-) | - | - | - |

**(b) Claude-3-opus after cross-critique**

| Variable Category | Responses* | | Accuracy | | Precision | | Recall | | | F1 score | |
| --- | --- | --- | --- | --- | --- | --- | --- | --- | --- | --- | --- |
|  | **Development** | **Test** | **Development** | **Test** | **Development** | **Test** | **Development** | **Test** | **Development** | | **Test** |
| Overall Results | 2 | 24 | 0.00 (-) | 0.45 (0.29-0.61) | 0.00 (-) | 0.46 (0.30-0.62) | - | 0.75 (0.59-0.91) | - | | 0.57 (0.41-0.73) |
| Trial Characteristics | 0 | 1 | - | 1.00 (-) | - | 1.00 (-) | - | 1.00 (-) | - | | 1.00 (-) |
| Population Characteristics | 0 | 16 | - | 0.53 (0.33-0.73) | - | 0.56 (0.36-0.76) | - | 0.81 (0.66-0.96) | - | | 0.66 (0.51-0.81) |
| Meta-analysis Outcomes | 2 | 7 | 0.00 (-) | 0.3 (0.00-0.67) | 0.00 (-) | 0.3 (0.00-0.67) | - | 0.50 (0.00-1.00) | - | | 0.38 (0.00-0.82) |

*Number of discordant responses generated for each variable category across all publications after cross-critique

Mean values with corresponding 95% confidence intervals for accuracy, precision, recall, and F1 score of (a) GPT-turbo and (b) Claude-3-opus for the subset of responses where GPT-4-turbo and Claude-3-opus generated discordant responses after cross-critique

### **Supplementary Table 9: Sensitivity analysis showing mean evaluation metrics with 95% confidence intervals for responses across all clinical trial publications included exclusively in the test set**

**(a) GPT-4-turbo in a zero-shot setting**

| Variable Category | Accuracy | Precision | Recall | F1 score |
| --- | --- | --- | --- | --- |
| Overall Results | 0.89 (0.88-0.90) | 0.98 (0.97-0.99) | 0.90 (0.89-0.91) | 0.93 (0.92-0.94) |
| Trial Characteristics | 1.00 (-) | 1.00 (-) | 1.00 (-) | 1.00 (-) |
| Population Characteristics | 0.84 (0.80-0.88) | 0.97 (0.95-0.99) | 0.85 (0.80-0.90) | 0.89 (0.86-0.92) |
| Meta-analysis Outcomes | 0.87 (0.79-0.95) | 1.00 (-) | 0.87 (0.79-0.95) | 0.90 (0.83-0.97) |

**(b) Claude-3-opus in a zero-shot setting**

| Variable Category | Accuracy | Precision | Recall | F1 score |
| --- | --- | --- | --- | --- |
| Overall Results | 0.92 (0.91-0.93) | 0.94 (0.93-0.95) | 0.95 (0.94-0.96) | 0.94 (0.93-0.95) |
| Trial Characteristics | 0.98 (0.97-0.99) | 1.00 (-) | 0.98 (0.97-0.99) | 0.99 (0.98-1.00) |
| Population Characteristics | 0.88 (0.85-0.91) | 0.88 (0.84-0.92) | 0.91 (0.88-0.94) | 0.89 (0.86-0.92) |
| Meta-analysis Outcomes | 0.91 (0.87-0.95) | 0.96 (0.92-1.00) | 0.96 (0.09) | 0.95 (0.93-0.99) |

Sensitivity analysis showing mean values with corresponding 95% confidence intervals for accuracy, precision, recall, and F1 score of (a) GPT-4-turbo and (b) Claude 3-opus for the responses of all publications of clinical trials included exclusively in the test set in a zero-shot setting

### **Supplementary Table 10: Sensitivity analysis showing mean evaluation metrics with 95% confidence intervals for responses with concordance across clinical trial publications included exclusively in the test set**

| Variable Category | Responses* | Accuracy | Precision | Recall | F1 score |
| --- | --- | --- | --- | --- | --- |
| Overall Results | 175 | 0.97 (0.96-0.98) | 1.00 (-) | 0.97 (0.96-0.98) | 0.98 (0.975-0.984) |
| Trial Characteristics | 61 | 1.00 (-) | 1.00 (-) | 1.00 (-) | 1.00 (-) |
| Population Characteristics | 75 | 0.96 (0.94-0.98) | 1.00 (-) | 0.95 (0.93-0.97) | 0.97 (0.95-0.99) |
| Meta-analysis Outcomes | 39 | 0.92 (0.86-0.98) | 1.00 (-) | 0.92 (0.87-0.97) | 0.95 (0.92-0.98) |

*Number of concordant responses generated for each variable category across publications of clinical trials included exclusively in the test set

Sensitivity analysis showing mean values with corresponding 95% confidence intervals for accuracy, precision, recall, and F1 score for the subset of responses where GPT-4-turbo and Claude-3-opus generated concordant responses across publications of clinical trials included exclusively in the test set

### **Supplementary Table 11: Sensitivity analysis showing mean evaluation metrics with 95% confidence intervals for responses with discordance across clinical trial publications included exclusively in the test set**

**(a) GPT-4-turbo in a zero-shot setting**

| Variable Category | Responses* | Accuracy | Precision | Recall | F1 score |
| --- | --- | --- | --- | --- | --- |
| Overall Results | 32 | 0.60 (0.43-0.77) | 0.57 (0.37-0.77) | 0.48 (0.30-0.67) | 0.51 (0.32-0.69) |
| Trial Characteristics | 2 | 1.00 (-) | 1.00 (-) | 1.00 (-) | 1.00 (-) |
| Population Characteristics | 24 | 0.54 (0.33-0.74) | 0.80 (0.59-1.00) | 0.66 (0.44-0.88) | 0.69 (0.48-0.90) |
| Meta-analysis Outcomes | 6 | 0.33 (0.00-0.80) | - | - | - |

**(b) Claude-3-opus in a zero-shot setting**

| Variable Category | Responses* | Accuracy | Precision | Recall | F1 score |
| --- | --- | --- | --- | --- | --- |
| Overall Results | 32 | 0.44 (0.00-0.18) | 0.44 (0.26-0.63) | 0.67 (0.49-0.85) | 0.53 (0.34-0.72) |
| Trial Characteristics | 2 | 0.50 (0.00-1.00) | 0.50 (0.00-1.00) | 0.50 (0.00-1.00) | 0.50 (0.00-1.00) |
| Population Characteristics | 24 | 0.50 (0.29-0.71) | 0.57 (0.36-0.79) | 0.67 (0.46-0.87) | 0.62 (0.40-0.83) |
| Meta-analysis Outcomes | 6 | 0.67 (0.20-1.00) | - | - | - |

*Number of discordant responses generated for each variable category across publications of clinical trials included exclusively in the test set

Sensitivity analysis showing mean values with corresponding 95% confidence intervals for accuracy, precision, recall, and F1 score of (a) GPT-4-turbo and (b) Claude 3-opus for a subset of responses where GPT-4-turbo and Claude-3-opus generated discordant responses across publications of clinical trials included exclusively in the test set

### **Supplementary Table 12: Sensitivity analysis showing mean evaluation metrics with 95% confidence intervals for responses with concordance after cross-critique across clinical trial publications included exclusively in the test set**

| Variable Category | Responses* | Accuracy | Precision | Recall | F1 score |
| --- | --- | --- | --- | --- | --- |
| Overall Results | 20 | 0.85 (0.75-0.95) | 0.78 (0.59-0.97) | 0.63 (0.45-0.81) | 0.70 (0.52-0.88) |
| Trial Characteristics | 4 | 0.83 (0.56-1.00) | 1.00 (-) | 0.83 (0.55-1.00) | 0.89 (0.70-1.00) |
| Population Characteristics | 14 | 0.79 (0.59-0.99) | 0.83 (0.62-1.00) | 0.64 (0.39-0.89) | 0.73 (0.50-0.96) |
| Meta-analysis Outcomes | 2 | 1.00 (-) | 0.50 (0.00-1.00) | 0.50 (0.00-1.00) | 0.50 (0.00-1.00) |

*Number of concordant responses generated for each variable category across publications of clinical trials included exclusively in the test set after cross-critique

Mean values with corresponding 95% confidence intervals for accuracy, precision, recall, and F1 score for the subset of responses where GPT-4-turbo and Claude-3-opus generated concordant responses after cross-critique across publications of clinical trials included exclusively in the test set

### **Supplementary Table 13: Sensitivity analysis showing mean evaluation metrics with 95% confidence intervals for responses with discordance after cross-critique across clinical trial publications included exclusively in the test set**

**(a) GPT-4-turbo after cross-critique**

| Variable Category | Responses* | Accuracy | Precision | Recall | F1 score |
| --- | --- | --- | --- | --- | --- |
| Overall Results | 12 | 0.15 (0.02-0.28) | 0.38 (0.10-0.65) | 0.29 (0.10-0.49) | 0.33 (0.10-0.56) |
| Trial Characteristics | 0 | - | - | - | - |
| Population Characteristics | 8 | 0.20 (0.01-0.39) | 0.67 (0.27-1.00) | 0.25 (0.05-0.45) | 0.36 (0.08-0.63) |
| Meta-analysis Outcomes | 4 | 0.17 (0.00-0.39) | 0.17 (0.00-0.39) | 0.50 (0.00-1.00) | 0.25 (0.00-0.59) |

**(b) Claude-3-opus after cross-critique**

| Variable Category | Responses* | Accuracy | Precision | Recall | F1 score |
| --- | --- | --- | --- | --- | --- |
| Overall Results | 12 | 0.55 (0.29-0.81) | 0.42 (0.14-0.69) | 0.47 (0.17-0.76) | 0.44 (0.15-0.73) |
| Trial Characteristics | 0 | - | - | - | - |
| Population Characteristics | 8 | 0.60 (0.31-0.89) | 0.50 (0.15-0.85) | 0.63 (0.29-0.96) | 0.56 (0.20-0.90) |
| Meta-analysis Outcomes | 4 | 0.50 (0.00-1.00) | 0.50 (0.00-1.00) | 0.50 (0.00-1.00) | 0.50 (0.00-1.00) |

*Number of discordant responses generated for each variable category across publications of clinical trials included exclusively in the test set after cross-critique

Mean values with corresponding 95% confidence intervals for accuracy, precision, recall, and F1 score of (a) GPT-turbo and (b) Claude-3-opus for the subset of responses where GPT-4-turbo and Claude-3-opus generated discordant responses after cross-critique across publications of clinical trials included exclusively in the test set

### **Supplementary Table 14: Mean evaluation metrics for responses by publications**

**(a) GPT-4-turbo (Prompt Development Set)**

| Trial | Variable Category | Accuracy | Precision | Recall | F1 score |
| --- | --- | --- | --- | --- | --- |
| *STAMPEDE 2019*  *(Clarke et al.)* | **Overall Results** | 1.00 | 1.00 | 1.00 | 1.00 |
|  | **Trial Characteristics** | 1.00 | 1.00 | 1.00 | 1.00 |
|  | **Population Characteristics** | 1.00 | 1.00 | 1.00 | 1.00 |
|  | **Meta-analysis Outcomes** | 1.00 | 1.00 | 1.00 | 1.00 |
| *ENZAMET 2023*  *(Sweeney et al.)* | **Overall Results** | 0.91 | 1.00 | 0.91 | 0.95 |
|  | **Trial Characteristics** | 1.00 | 1.00 | 1.00 | 1.00 |
|  | **Population Characteristics** | 1.00 | 1.00 | 1.00 | 1.00 |
|  | **Meta-analysis Outcomes** | 0.60 | 1.00 | 0.60 | 0.75 |
| *LATITUDE 2017*  *(Fizazi et al.)* | **Overall Results** | 0.91 | 1.00 | 0.82 | 0.90 |
|  | **Trial Characteristics** | 1.00 | 1.00 | 1.00 | 1.00 |
|  | **Population Characteristics** | 0.82 | 1.00 | 0.33 | 0.50 |
|  | **Meta-analysis Outcomes** | 1.00 | 1.00 | 1.00 | 1.00 |
| *LATITUDE 2019*  *(Fizazi et al.)* | **Overall Results** | 0.91 | 1.00 | 0.89 | 0.94 |
|  | **Trial Characteristics** | 1.00 | 1.00 | 1.00 | 1.00 |
|  | **Population Characteristics** | 0.82 | 1.00 | 0.78 | 0.88 |
|  | **Meta-analysis Outcomes** | 1.00 | 1.00 | 1.00 | 1.00 |
| *CHAARTED 2015*  *(Sweeney et al.)* | **Overall Results** | 1.00 | 1.00 | 1.00 | 1.00 |
|  | **Trial Characteristics** | 1.00 | 1.00 | 1.00 | 1.00 |
|  | **Population Characteristics** | 1.00 | 1.00 | 1.00 | 1.00 |
|  | **Meta-analysis Outcomes** | 1.00 | 1.00 | 1.00 | 1.00 |

**(b) GPT-4-turbo (Test Set)**

| Trial | Variable Category | Accuracy | Precision | Recall | F1 score |
| --- | --- | --- | --- | --- | --- |
| *GETUG 2015*  *(Gravis et al.)* | **Overall Results** | 0.96 | 0.96 | 1.00 | 0.98 |
|  | **Trial Characteristics** | 1.00 | 1.00 | 1.00 | 1.00 |
|  | **Population Characteristics** | 0.91 | 0.91 | 1.00 | 0.95 |
|  | **Meta-analysis Outcomes** | 1.00 | 1.00 | 1.00 | 1.00 |
| *GETUG 2013*  *(Gravis et al.)* | **Overall Results** | 1.00 | 1.00 | 1.00 | 1.00 |
|  | **Trial Characteristics** | 1.00 | 1.00 | 1.00 | 1.00 |
|  | **Population Characteristics** | 1.00 | 1.00 | 1.00 | 1.00 |
|  | **Meta-analysis Outcomes** | 1.00 | 1.00 | 1.00 | 1.00 |
| *STAMPEDE 2023*  *(Attard et al.)* | **Overall Results** | 1.00 | 1.00 | 1.00 | 1.00 |
|  | **Trial Characteristics** | 1.00 | 1.00 | 1.00 | 1.00 |
|  | **Population Characteristics** | 1.00 | 1.00 | 1.00 | 1.00 |
|  | **Meta-analysis Outcomes** | 1.00 | 1.00 | 1.00 | 1.00 |
| *STAMPEDE 2019*  *(Hoyle et al.)* | **Overall Results** | 0.91 | 0.94 | 0.94 | 0.94 |
|  | **Trial Characteristics** | 1.00 | 1.00 | 1.00 | 1.00 |
|  | **Population Characteristics** | 0.91 | 0.90 | 1.00 | 0.95 |
|  | **Meta-analysis Outcomes** | 0.80 | 1.00 | 0.67 | 0.80 |
| *STAMPEDE 2022*  *(James et al.)* | **Overall Results** | 0.91 | 0.88 | 1.00 | 0.93 |
|  | **Trial Characteristics** | 1.00 | 1.00 | 1.00 | 1.00 |
|  | **Population Characteristics** | 0.82 | 0.78 | 1.00 | 0.88 |
|  | **Meta-analysis Outcomes** | 1.00 | 1.00 | 1.00 | 1.00 |
| *STAMPEDE 2016*  *(James et al.)* | **Overall Results** | 0.78 | 1.00 | 0.71 | 0.83 |
|  | **Trial Characteristics** | 1.00 | 1.00 | 1.00 | 1.00 |
|  | **Population Characteristics** | 0.73 | 1.00 | 0.57 | 0.72 |
|  | **Meta-analysis Outcomes** | 0.60 | 1.00 | 0.33 | 0.50 |
| *STAMPEDE 2017*  *(James et al.)* | **Overall Results** | 0.74 | 1.00 | 0.65 | 0.79 |
|  | **Trial Characteristics** | 0.86 | 1.00 | 0.86 | 0.92 |
|  | **Population Characteristics** | 0.73 | 1.00 | 0.57 | 0.73 |
|  | **Meta-analysis Outcomes** | 0.60 | 1.00 | 0.33 | 0.50 |
| *STAMPEDE 2018*  *(Sydes et al.)* | **Overall Results** | 0.78 | 0.92 | 0.75 | 0.83 |
|  | **Trial Characteristics** | 1.00 | 1.00 | 1.00 | 1.00 |
|  | **Population Characteristics** | 0.73 | 0.80 | 0.67 | 0.73 |
|  | **Meta-analysis Outcomes** | 0.60 | 1.00 | 0.33 | 0.50 |
| *CHAARTED 2018*  *(Kyriakopoulos*  *et al.)* | **Overall Results** | 0.96 | 1.00 | 0.95 | 0.97 |
|  | **Trial Characteristics** | 1.00 | 1.00 | 1.00 | 1.00 |
|  | **Population Characteristics** | 0.91 | 1.00 | 0.89 | 0.94 |
|  | **Meta-analysis Outcomes** | 1.00 | 1.00 | 1.00 | 1.00 |
| *SWOG 2022*  *(Agarwal et al.)* | **Overall Results** | 1.00 | 1.00 | 1.00 | 1.00 |
|  | **Trial Characteristics** | 1.00 | 1.00 | 1.00 | 1.00 |
|  | **Population Characteristics** | 1.00 | 1.00 | 1.00 | 1.00 |
|  | **Meta-analysis Outcomes** | 1.00 | 1.00 | 1.00 | 1.00 |
| *PEACE 1 2022*  *(Fizazi et al.)* | **Overall Results** | 0.78 | 1.00 | 0.75 | 0.86 |
|  | **Trial Characteristics** | 1.00 | 1.00 | 1.00 | 1.00 |
|  | **Population Characteristics** | 0.55 | 1.00 | 0.50 | 0.67 |
|  | **Meta-analysis Outcomes** | 1.00 | 1.00 | 1.00 | 1.00 |
| *ENZAMET 2019*  *(Davis et al.)* | **Overall Results** | 0.87 | 0.94 | 0.89 | 0.91 |
|  | **Trial Characteristics** | 1.00 | 1.00 | 1.00 | 1.00 |
|  | **Population Characteristics** | 0.82 | 1.00 | 0.78 | 0.88 |
|  | **Meta-analysis Outcomes** | 0.80 | 0.67 | 1.00 | 0.80 |
| *TITAN 2021*  *(Chi et al.)* | **Overall Results** | 0.70 | 1.00 | 0.67 | 0.80 |
|  | **Trial Characteristics** | 1.00 | 1.00 | 1.00 | 1.00 |
|  | **Population Characteristics** | 0.55 | 1.00 | 0.44 | 0.62 |
|  | **Meta-analysis Outcomes** | 0.60 | 1.00 | 0.60 | 0.75 |
| *TITAN 2019*  *(Chi et al.)* | **Overall Results** | 0.65 | 0.88 | 0.71 | 0.79 |
|  | **Trial Characteristics** | 1.00 | 1.00 | 1.00 | 1.00 |
|  | **Population Characteristics** | 0.64 | 0.78 | 0.78 | 0.78 |
|  | **Meta-analysis Outcomes** | 0.20 | 1.00 | 0.20 | 0.33 |
| *ARCHES 2022*  *(Armstrong et al.)* | **Overall Results** | 0.96 | 1.00 | 0.95 | 0.98 |
|  | **Trial Characteristics** | 1.00 | 1.00 | 1.00 | 1.00 |
|  | **Population Characteristics** | 0.91 | 1.00 | 0.91 | 0.95 |
|  | **Meta-analysis Outcomes** | 1.00 | 1.00 | 1.00 | 1.00 |
| *ARASENS 2023*  *(Hussain et al.)* | **Overall Results** | 1.00 | 1.00 | 1.00 | 1.00 |
|  | **Trial Characteristics** | 1.00 | 1.00 | 1.00 | 1.00 |
|  | **Population Characteristics** | 1.00 | 1.00 | 1.00 | 1.00 |
|  | **Meta-analysis Outcomes** | 1.00 | 1.00 | 1.00 | 1.00 |
| *ARASENS 2022*  *(Smith et al.)* | **Overall Results** | 1.00 | 1.00 | 1.00 | 1.00 |
|  | **Trial Characteristics** | 1.00 | 1.00 | 1.00 | 1.00 |
|  | **Population Characteristics** | 1.00 | 1.00 | 1.00 | 1.00 |
|  | **Meta-analysis Outcomes** | 1.00 | 1.00 | 1.00 | 1.00 |

**(c) Claude-3-opus (Prompt Development Set)**

| Trial | Variable Category | Accuracy | Precision | Recall | F1 score |
| --- | --- | --- | --- | --- | --- |
| *STAMPEDE 2019*  *(Clarke et al.)* | **Overall Results** | 0.96 | 0.94 | 1.00 | 0.97 |
|  | **Trial Characteristics** | 1.00 | 1.00 | 1.00 | 1.00 |
|  | **Population Characteristics** | 1.00 | 1.00 | 1.00 | 1.00 |
|  | **Meta-analysis Outcomes** | 0.80 | 0.75 | 1.00 | 0.86 |
| *ENZAMET 2023*  *(Sweeney et al.)* | **Overall Results** | 0.91 | 1.00 | 1.00 | 0.95 |
|  | **Trial Characteristics** | 1.00 | 1.00 | 1.00 | 1.00 |
|  | **Population Characteristics** | 1.00 | 1.00 | 1.00 | 1.00 |
|  | **Meta-analysis Outcomes** | 0.60 | 0.60 | 1.00 | 0.75 |
| *LATITUDE 2017*  *(Fizazi et al.)* | **Overall Results** | 0.96 | 1.00 | 0.91 | 0.95 |
|  | **Trial Characteristics** | 1.00 | 1.00 | 1.00 | 1.00 |
|  | **Population Characteristics** | 0.91 | 1.00 | 0.67 | 0.80 |
|  | **Meta-analysis Outcomes** | 1.00 | 1.00 | 1.00 | 1.00 |
| *LATITUDE 2019*  *(Fizazi et al.)* | **Overall Results** | 0.96 | 1.00 | 0.89 | 0.94 |
|  | **Trial Characteristics** | 1.00 | 1.00 | 1.00 | 1.00 |
|  | **Population Characteristics** | 0.91 | 1.00 | 0.89 | 0.94 |
|  | **Meta-analysis Outcomes** | 1.00 | 1.00 | 1.00 | 1.00 |
| *CHAARTED 2015*  *(Sweeney et al.)* | **Overall Results** | 1.00 | 1.00 | 1.00 | 1.00 |
|  | **Trial Characteristics** | 1.00 | 1.00 | 1.00 | 1.00 |
|  | **Population Characteristics** | 1.00 | 1.00 | 1.00 | 1.00 |
|  | **Meta-analysis Outcomes** | 1.00 | 1.00 | 1.00 | 1.00 |

**(d) Claude-3-opus (Test Set)**

| Trial | Variable Category | Accuracy | Precision | Recall | F1 score |
| --- | --- | --- | --- | --- | --- |
| *GETUG 2015*  *(Gravis et al.)* | **Overall Results** | 1.00 | 1.00 | 1.00 | 1.00 |
|  | **Trial Characteristics** | 1.00 | 1.00 | 1.00 | 1.00 |
|  | **Population Characteristics** | 1.00 | 1.00 | 1.00 | 1.00 |
|  | **Meta-analysis Outcomes** | 1.00 | 1.00 | 1.00 | 1.00 |
| *GETUG 2013*  *(Gravis et al.)* | **Overall Results** | 0.96 | 0.93 | 1.00 | 0.97 |
|  | **Trial Characteristics** | 1.00 | 1.00 | 1.00 | 1.00 |
|  | **Population Characteristics** | 0.91 | 0.86 | 1.00 | 0.92 |
|  | **Meta-analysis Outcomes** | 1.00 | 1.00 | 1.00 | 1.00 |
| *STAMPEDE 2023*  *(Attard et al.)* | **Overall Results** | 1.00 | 1.00 | 1.00 | 1.00 |
|  | **Trial Characteristics** | 1.00 | 1.00 | 1.00 | 1.00 |
|  | **Population Characteristics** | 1.00 | 1.00 | 1.00 | 1.00 |
|  | **Meta-analysis Outcomes** | 1.00 | 1.00 | 1.00 | 1.00 |
| *STAMPEDE 2019*  *(Hoyle et al.)* | **Overall Results** | 0.87 | 0.85 | 1.00 | 0.92 |
|  | **Trial Characteristics** | 1.00 | 1.00 | 1.00 | 1.00 |
|  | **Population Characteristics** | 1.00 | 1.00 | 1.00 | 1.00 |
|  | **Meta-analysis Outcomes** | 0.40 | 0.25 | 1.00 | 0.40 |
| *STAMPEDE 2022*  *(James et al.)* | **Overall Results** | 0.91 | 1.00 | 0.88 | 0.93 |
|  | **Trial Characteristics** | 1.00 | 1.00 | 1.00 | 1.00 |
|  | **Population Characteristics** | 0.82 | 1.00 | 0.78 | 0.88 |
|  | **Meta-analysis Outcomes** | 1.00 | 1.00 | 1.00 | 1.00 |
| *STAMPEDE 2016*  *(James et al.)* | **Overall Results** | 0.74 | 0.73 | 0.85 | 0.79 |
|  | **Trial Characteristics** | 1.00 | 1.00 | 1.00 | 1.00 |
|  | **Population Characteristics** | 0.64 | 0.43 | 1.00 | 0.60 |
|  | **Meta-analysis Outcomes** | 0.60 | 1.00 | 0.33 | 0.50 |
| *STAMPEDE 2017*  *(James et al.)* | **Overall Results** | 0.83 | 1.00 | 0.76 | 0.79 |
|  | **Trial Characteristics** | 1.00 | 1.00 | 1.00 | 1.00 |
|  | **Population Characteristics** | 0.82 | 1.00 | 0.71 | 0.83 |
|  | **Meta-analysis Outcomes** | 0.60 | 1.00 | 0.33 | 0.50 |
| *STAMPEDE 2018*  *(Sydes et al.)* | **Overall Results** | 0.78 | 1.00 | 0.71 | 0.83 |
|  | **Trial Characteristics** | 1.00 | 1.00 | 1.00 | 1.00 |
|  | **Population Characteristics** | 0.73 | 1.00 | 0.57 | 0.73 |
|  | **Meta-analysis Outcomes** | 0.60 | 1.00 | 0.33 | 0.50 |
| *CHAARTED 2018*  *(Kyriakopoulos*  *et al.)* | **Overall Results** | 1.00 | 1.00 | 1.00 | 1.00 |
|  | **Trial Characteristics** | 1.00 | 1.00 | 1.00 | 1.00 |
|  | **Population Characteristics** | 1.00 | 1.00 | 1.00 | 1.00 |
|  | **Meta-analysis Outcomes** | 1.00 | 1.00 | 1.00 | 1.00 |
| *SWOG 2022*  *(Agarwal et al.)* | **Overall Results** | 0.91 | 0.85 | 1.00 | 0.92 |
|  | **Trial Characteristics** | 1.00 | 1.00 | 1.00 | 1.00 |
|  | **Population Characteristics** | 0.82 | 0.60 | 1.00 | 0.75 |
|  | **Meta-analysis Outcomes** | 1.00 | 1.00 | 1.00 | 1.00 |
| *PEACE 1 2022*  *(Fizazi et al.)* | **Overall Results** | 1.00 | 1.00 | 1.00 | 1.00 |
|  | **Trial Characteristics** | 1.00 | 1.00 | 1.00 | 1.00 |
|  | **Population Characteristics** | 1.00 | 1.00 | 1.00 | 1.00 |
|  | **Meta-analysis Outcomes** | 1.00 | 1.00 | 1.00 | 1.00 |
| *ENZAMET 2019*  *(Davis et al.)* | **Overall Results** | 0.87 | 0.94 | 0.89 | 0.91 |
|  | **Trial Characteristics** | 1.00 | 1.00 | 1.00 | 1.00 |
|  | **Population Characteristics** | 0.82 | 1.00 | 0.78 | 0.88 |
|  | **Meta-analysis Outcomes** | 0.80 | 0.67 | 1.00 | 0.80 |
| *TITAN 2021*  *(Chi et al.)* | **Overall Results** | 0.87 | 1.00 | 0.86 | 0.92 |
|  | **Trial Characteristics** | 1.00 | 1.00 | 1.00 | 1.00 |
|  | **Population Characteristics** | 0.82 | 1.00 | 0.78 | 0.88 |
|  | **Meta-analysis Outcomes** | 0.80 | 1.00 | 0.80 | 0.89 |
| *TITAN 2019*  *(Chi et al.)* | **Overall Results** | 0.96 | 1.00 | 0.96 | 0.98 |
|  | **Trial Characteristics** | 1.00 | 1.00 | 1.00 | 1.00 |
|  | **Population Characteristics** | 1.00 | 1.00 | 1.00 | 1.00 |
|  | **Meta-analysis Outcomes** | 0.80 | 1.00 | 0.80 | 0.89 |
| *ARCHES 2022*  *(Armstrong et al.)* | **Overall Results** | 0.87 | 1.00 | 0.86 | 0.92 |
|  | **Trial Characteristics** | 0.86 | 1.00 | 0.86 | 0.92 |
|  | **Population Characteristics** | 0.82 | 1.00 | 0.81 | 0.90 |
|  | **Meta-analysis Outcomes** | 1.00 | 1.00 | 1.00 | 1.00 |
| *ARASENS 2023*  *(Hussain et al.)* | **Overall Results** | 0.91 | 0.91 | 1.00 | 0.95 |
|  | **Trial Characteristics** | 1.00 | 1.00 | 1.00 | 1.00 |
|  | **Population Characteristics** | 1.00 | 1.00 | 1.00 | 1.00 |
|  | **Meta-analysis Outcomes** | 0.60 | 0.60 | 1.00 | 0.75 |
| *ARASENS 2022*  *(Smith et al.)* | **Overall Results** | 0.78 | 0.79 | 0.85 | 0.81 |
|  | **Trial Characteristics** | 1.00 | 1.00 | 1.00 | 1.00 |
|  | **Population Characteristics** | 0.54 | 0.50 | 0.60 | 0.55 |
|  | **Meta-analysis Outcomes** | 1.00 | 1.00 | 1.00 | 1.00 |

Mean values for accuracy, precision, recall, and F1 scores of (a) GPT-4-turbo in prompt development set (b) GPT-4-turbo in test set (c) Claude-3-opus in prompt development set, and (d) Claude-3-opus in test set for all variable categories by publications

### **Supplementary Table 15: Mean evaluation metrics for the subset of responses with concordance by publications**

**(a) Prompt Development Set**

| Trial | Variable Category | Responses* | Accuracy | Precision | Recall | F1 score |
| --- | --- | --- | --- | --- | --- | --- |
| *STAMPEDE 2019*  *(Clarke et al.)* | **Overall Results** | 22 | 1.00 | 1.00 | 1.00 | 1.00 |
|  | **Trial Characteristics** | 7 | 1.00 | 1.00 | 1.00 | 1.00 |
|  | **Population Characteristics** | 11 | 1.00 | 1.00 | 1.00 | 1.00 |
|  | **Meta-analysis Outcomes** | 4 | 1.00 | 1.00 | 1.00 | 1.00 |
| *ENZAMET 2023*  *(Sweeney et al.)* | **Overall Results** | 21 | 1.00 | 1.00 | 1.00 | 1.00 |
|  | **Trial Characteristics** | 7 | 1.00 | 1.00 | 1.00 | 1.00 |
|  | **Population Characteristics** | 11 | 1.00 | 1.00 | 1.00 | 1.00 |
|  | **Meta-analysis Outcomes** | 3 | 1.00 | 1.00 | 1.00 | 1.00 |
| *LATITUDE 2017*  *(Fizazi et al.)* | **Overall Results** | 22 | 0.95 | 1.00 | 0.90 | 0.95 |
|  | **Trial Characteristics** | 7 | 1.00 | 1.00 | 1.00 | 1.00 |
|  | **Population Characteristics** | 10 | 0.90 | 1.00 | 0.50 | 0.67 |
|  | **Meta-analysis Outcomes** | 5 | 1.00 | 1.00 | 1.00 | 1.00 |
| *LATITUDE 2019*  *(Fizazi et al.)* | **Overall Results** | 22 | 0.95 | 1.00 | 0.94 | 0.97 |
|  | **Trial Characteristics** | 7 | 1.00 | 1.00 | 1.00 | 1.00 |
|  | **Population Characteristics** | 10 | 0.90 | 1.00 | 0.88 | 0.93 |
|  | **Meta-analysis Outcomes** | 5 | 1.00 | 1.00 | 1.00 | 1.00 |
| *CHAARTED 2015*  *(Sweeney et al.)* | **Overall Results** | 23 | 1.00 | 1.00 | 1.00 | 1.00 |
|  | **Trial Characteristics** | 7 | 1.00 | 1.00 | 1.00 | 1.00 |
|  | **Population Characteristics** | 11 | 1.00 | 1.00 | 1.00 | 1.00 |
|  | **Meta-analysis Outcomes** | 5 | 1.00 | 1.00 | 1.00 | 1.00 |

**(b) Test Set**

| Trial | Variable Category | Responses* | Accuracy | Precision | Recall | F1 score |
| --- | --- | --- | --- | --- | --- | --- |
| *GETUG 2015*  *(Gravis et al.)* | **Overall Results** | 22 | 1.00 | 1.00 | 1.00 | 1.00 |
|  | **Trial Characteristics** | 7 | 1.00 | 1.00 | 1.00 | 1.00 |
|  | **Population Characteristics** | 10 | 1.00 | 1.00 | 1.00 | 1.00 |
|  | **Meta-analysis Outcomes** | 5 | 1.00 | 1.00 | 1.00 | 1.00 |
| *GETUG 2013*  *(Gravis et al.)* | **Overall Results** | 22 | 1.00 | 1.00 | 1.00 | 1.00 |
|  | **Trial Characteristics** | 7 | 1.00 | 1.00 | 1.00 | 1.00 |
|  | **Population Characteristics** | 10 | 1.00 | 1.00 | 1.00 | 1.00 |
|  | **Meta-analysis Outcomes** | 5 | 1.00 | 1.00 | 1.00 | 1.00 |
| *STAMPEDE 2023*  *(Attard et al.)* | **Overall Results** | 23 | 1.00 | 1.00 | 1.00 | 1.00 |
|  | **Trial Characteristics** | 7 | 1.00 | 1.00 | 1.00 | 1.00 |
|  | **Population Characteristics** | 11 | 1.00 | 1.00 | 1.00 | 1.00 |
|  | **Meta-analysis Outcomes** | 5 | 1.00 | 1.00 | 1.00 | 1.00 |
| *STAMPEDE 2019*  *(Hoyle et al.)* | **Overall Results** | 19 | 1.00 | 1.00 | 1.00 | 1.00 |
|  | **Trial Characteristics** | 7 | 1.00 | 1.00 | 1.00 | 1.00 |
|  | **Population Characteristics** | 10 | 1.00 | 1.00 | 1.00 | 1.00 |
|  | **Meta-analysis Outcomes** | 2 | 1.00 | 1.00 | 1.00 | 1.00 |
| *STAMPEDE 2022*  *(James et al.)* | **Overall Results** | 21 | 1.00 | 1.00 | 1.00 | 1.00 |
|  | **Trial Characteristics** | 7 | 1.00 | 1.00 | 1.00 | 1.00 |
|  | **Population Characteristics** | 9 | 1.00 | 1.00 | 1.00 | 1.00 |
|  | **Meta-analysis Outcomes** | 5 | 1.00 | 1.00 | 1.00 | 1.00 |
| *STAMPEDE 2016*  *(James et al.)* | **Overall Results** | 18 | 0.89 | 1.00 | 0.83 | 0.91 |
|  | **Trial Characteristics** | 7 | 1.00 | 1.00 | 1.00 | 1.00 |
|  | **Population Characteristics** | 6 | 1.00 | 1.00 | 1.00 | 1.00 |
|  | **Meta-analysis Outcomes** | 5 | 0.60 | 1.00 | 0.33 | 0.50 |
| *STAMPEDE 2017*  *(James et al.)* | **Overall Results** | 21 | 0.81 | 1.00 | 0.73 | 0.85 |
|  | **Trial Characteristics** | 6 | 1.00 | 1.00 | 1.00 | 1.00 |
|  | **Population Characteristics** | 10 | 0.80 | 1.00 | 0.67 | 0.80 |
|  | **Meta-analysis Outcomes** | 5 | 0.60 | 1.00 | 0.33 | 0.50 |
| *STAMPEDE 2018*  *(Sydes et al.)* | **Overall Results** | 21 | 0.81 | 1.00 | 0.73 | 0.85 |
|  | **Trial Characteristics** | 7 | 1.00 | 1.00 | 1.00 | 1.00 |
|  | **Population Characteristics** | 9 | 0.78 | 1.00 | 0.60 | 0.75 |
|  | **Meta-analysis Outcomes** | 5 | 0.60 | 1.00 | 0.33 | 0.50 |
| *CHAARTED 2018*  *(Kyriakopoulos*  *et al.)* | **Overall Results** | 22 | 1.00 | 1.00 | 1.00 | 1.00 |
|  | **Trial Characteristics** | 7 | 1.00 | 1.00 | 1.00 | 1.00 |
|  | **Population Characteristics** | 10 | 1.00 | 1.00 | 1.00 | 1.00 |
|  | **Meta-analysis Outcomes** | 5 | 1.00 | 1.00 | 1.00 | 1.00 |
| *SWOG 2022*  *(Agarwal et al.)* | **Overall Results** | 21 | 1.00 | 1.00 | 1.00 | 1.00 |
|  | **Trial Characteristics** | 7 | 1.00 | 1.00 | 1.00 | 1.00 |
|  | **Population Characteristics** | 9 | 1.00 | 1.00 | 1.00 | 1.00 |
|  | **Meta-analysis Outcomes** | 5 | 1.00 | 1.00 | 1.00 | 1.00 |
| *PEACE 1 2022*  *(Fizazi et al.)* | **Overall Results** | 15 | 1.00 | 1.00 | 1.00 | 1.00 |
|  | **Trial Characteristics** | 6 | 1.00 | 1.00 | 1.00 | 1.00 |
|  | **Population Characteristics** | 4 | 1.00 | 1.00 | 1.00 | 1.00 |
|  | **Meta-analysis Outcomes** | 5 | 1.00 | 1.00 | 1.00 | 1.00 |
| *ENZAMET 2019*  *(Davis et al.)* | **Overall Results** | 22 | 0.91 | 1.00 | 0.89 | 0.94 |
|  | **Trial Characteristics** | 7 | 1.00 | 1.00 | 1.00 | 1.00 |
|  | **Population Characteristics** | 11 | 0.82 | 1.00 | 0.78 | 0.88 |
|  | **Meta-analysis Outcomes** | 4 | 1.00 | 1.00 | 1.00 | 1.00 |
| *TITAN 2021*  *(Chi et al.)* | **Overall Results** | 19 | 0.84 | 1.00 | 0.82 | 0.90 |
|  | **Trial Characteristics** | 7 | 1.00 | 1.00 | 1.00 | 1.00 |
|  | **Population Characteristics** | 8 | 0.75 | 1.00 | 0.67 | 0.80 |
|  | **Meta-analysis Outcomes** | 4 | 0.75 | 1.00 | 0.75 | 0.86 |
| *TITAN 2019*  *(Chi et al.)* | **Overall Results** | 16 | 0.94 | 1.00 | 0.94 | 0.97 |
|  | **Trial Characteristics** | 7 | 1.00 | 1.00 | 1.00 | 1.00 |
|  | **Population Characteristics** | 7 | 1.00 | 1.00 | 1.00 | 1.00 |
|  | **Meta-analysis Outcomes** | 2 | 0.50 | 1.00 | 0.50 | 0.67 |
| *ARCHES 2022*  *(Armstrong et al.)* | **Overall Results** | 21 | 0.95 | 1.00 | 0.95 | 0.97 |
|  | **Trial Characteristics** | 6 | 1.00 | 1.00 | 1.00 | 1.00 |
|  | **Population Characteristics** | 10 | 0.90 | 1.00 | 0.90 | 0.95 |
|  | **Meta-analysis Outcomes** | 5 | 1.00 | 1.00 | 1.00 | 1.00 |
| *ARASENS 2023*  *(Hussain et al.)* | **Overall Results** | 21 | 1.00 | 1.00 | 1.00 | 1.00 |
|  | **Trial Characteristics** | 7 | 1.00 | 1.00 | 1.00 | 1.00 |
|  | **Population Characteristics** | 11 | 1.00 | 1.00 | 1.00 | 1.00 |
|  | **Meta-analysis Outcomes** | 3 | 1.00 | 1.00 | 1.00 | 1.00 |
| *ARASENS 2022*  *(Smith et al.)* | **Overall Results** | 18 | 1.00 | 1.00 | 1.00 | 1.00 |
|  | **Trial Characteristics** | 7 | 1.00 | 1.00 | 1.00 | 1.00 |
|  | **Population Characteristics** | 6 | 1.00 | 1.00 | 1.00 | 1.00 |
|  | **Meta-analysis Outcomes** | 5 | 1.00 | 1.00 | 1.00 | 1.00 |

*Number of concordant responses generated for each variable category by publications

Mean values for accuracy, precision, recall, and F1 score of (a) development and (b) test sets for the subset of responses where GPT-4-turbo and Claude-3-opus generated concordant responses by publications

### **Supplementary Table 16: Mean evaluation metrics for a subset of responses with discordance by publications**

**(a) GPT-4-turbo (Prompt Development Set)**

| Trial | Variable Category | Responses* | Accuracy | Precision | Recall | F1 score |
| --- | --- | --- | --- | --- | --- | --- |
| *STAMPEDE 2019*  *(Clarke et al.)* | **Overall Results** | 1 | 1.00 | 1.00 | 1.00 | 1.00 |
|  | **Meta-analysis Outcomes** | 1 | 1.00 | 1.00 | 1.00 | 1.00 |
| *ENZAMET 2023*  *(Sweeney*  *et al.)* | **Overall Results** | 2 | 0.00 | - | 0.00 | - |
|  | **Meta-analysis Outcomes** | 2 | 0.00 | - | 0.00 | - |
| *LATITUDE 2017*  *(Fizazi et al.)* | **Overall Results** | 1 | 0.00 | - | 0.00 | - |
|  | **Population Characteristics** | 1 | 0.00 | - | 0.00 | - |
| *LATITUDE 2019*  *(Fizazi et al.)* | **Overall Results** | 1 | 0.00 | - | 0.00 | - |
|  | **Population Characteristics** | 1 | 0.00 | - | 0.00 | - |

**(b) GPT-4-turbo (Test Set)**

| Trial | Variable Category | Responses* | Accuracy | Precision | Recall | F1 score |
| --- | --- | --- | --- | --- | --- | --- |
| *GETUG 2015*  *(Gravis et al.)* | **Overall Results** | 1 | 0.00 | 0.00 | - | - |
|  | **Population Characteristics** | 1 | 0.00 | 0.00 | - | - |
| *GETUG 2013*  *(Gravis et al.)* | **Overall Results** | 1 | 1.00 | 1.00 | 1.00 | 1.00 |
|  | **Population Characteristics** | 1 | 1.00 | 1.00 | 1.00 | 1.00 |
| *STAMPEDE 2019*  *(Attard et al.)* | **Overall Results** | 4 | 0.50 | 0.50 | 0.50 | 0.50 |
|  | **Population Characteristics** | 1 | 0.00 | 0.00 | - | - |
|  | **Meta-analysis Outcomes** | 3 | 0.67 | 1.00 | 0.50 | 0.67 |
| *STAMPEDE 2022*  *(James et al.)* | **Overall Results** | 2 | 0.00 | 0.00 | - | - |
|  | **Population Characteristics** | 2 | 0.00 | 0.00 | - | - |
| *STAMPEDE 2016*  *(James et al.)* | **Overall Results** | 5 | 0.40 | 1.00 | 0.40 | 0.57 |
|  | **Population Characteristics** | 5 | 0.40 | 1.00 | 0.40 | 0.57 |
| *STAMPEDE 2017*  *(James et al.)* | **Overall Results** | 2 | 0.00 | 0.00 | 0.00 | 0.00 |
|  | **Trial Characteristics** | 1 | 0.00 | - | 0.00 | - |
|  | **Population Characteristics** | 1 | 0.00 | - | 0.00 | - |
| *STAMPEDE 2018*  *(Sydes et al.)* | **Overall Results** | 2 | 0.50 | 0.50 | 1.00 | 0.67 |
|  | **Population Characteristics** | 2 | 0.50 | 0.50 | 1.00 | 0.67 |
| *CHAARTED 2018*  *(Kyriakopoulos*  *et al.)* | **Overall Results** | 1 | 0.00 | - | 0.00 | - |
|  | **Population Characteristics** | 1 | 0.00 | - | 0.00 | - |
| *SWOG 2022*  *(Agarwal et al.)* | **Overall Results** | 2 | 1.00 | - | - | - |
|  | **Population Characteristics** | 2 | 1.00 | - | - | - |
| *PEACE 1 2022*  *(Fizazi et al.)* | **Overall Results** | 8 | 0.38 | 1.00 | 0.38 | 0.55 |
|  | **Trial Characteristics** | 1 | 1.00 | 1.00 | 1.00 | 1.00 |
|  | **Population Characteristics** | 7 | 0.29 | 1.00 | 0.29 | 0.44 |
| *ENZAMET 2019*  *(Davis et al.)* | **Overall Results** | 1 | 0.00 | 0.00 | - | - |
|  | **Meta-analysis Outcomes** | 1 | 0.00 | 0.00 | - | - |
| *TITAN 2021*  *(Chi et al.)* | **Overall Results** | 4 | 0.00 | 0.00 | 0.00 | 0.00 |
|  | **Population Characteristics** | 3 | 0.00 | - | 0.00 | - |
|  | **Meta-analysis Outcomes** | 1 | 0.00 | - | 0.00 | - |
| *TITAN 2019*  *(Chi et al.)* | **Overall Results** | 7 | 0.00 | 0.00 | 0.00 | 0.00 |
|  | **Population Characteristics** | 4 | 0.00 | 0.00 | 0.00 | - |
|  | **Meta-analysis Outcomes** | 3 | 0.00 | - | 0.00 | - |
| *ARCHES 2022*  *(Armstrong et al.)* | **Overall Results** | 2 | 1.00 | 1.00 | 1.00 | 1.00 |
|  | **Trial Characteristics** | 1 | 1.00 | 1.00 | 1.00 | 1.00 |
|  | **Population Characteristics** | 1 | 1.00 | 1.00 | 1.00 | 1.00 |
| *ARASENS 2023*  *(Hussain et al.)* | **Overall Results** | 2 | 1.00 | - | - | - |
|  | **Meta-analysis Outcomes** | 2 | 1.00 | - | - | - |
| *ARASENS 2022*  *(Smith et al.)* | **Overall Results** | 5 | 1.00 | 1.00 | 1.00 | 1.00 |
|  | **Population Characteristics** | 5 | 1.00 | 1.00 | 1.00 | 1.00 |

**(c) Claude-3-opus (Prompt Development Set)**

| Trial | Variable Category | Responses* | Accuracy | Precision | Recall | F1 score |
| --- | --- | --- | --- | --- | --- | --- |
| *STAMPEDE 2019*  *(Clarke et al.)* | **Overall Results** | 1 | 0.00 | 0.00 | - | - |
|  | **Meta-analysis Outcomes** | 1 | 0.00 | 0.00 | - | - |
| *ENZAMET 2023*  *(Sweeney*  *et al.)* | **Overall Results** | 2 | 0.00 | 0.00 | - | - |
|  | **Meta-analysis Outcomes** | 2 | 0.00 | 0.00 | - | - |
| *LATITUDE 2017*  *(Fizazi et al.)* | **Overall Results** | 1 | 1.00 | 1.00 | 1.00 | 1.00 |
|  | **Population Characteristics** | 1 | 1.00 | 1.00 | 1.00 | 1.00 |
| *LATITUDE 2019*  *(Fizazi et al.)* | **Overall Results** | 1 | 1.00 | 1.00 | 1.00 | 1.00 |
|  | **Population Characteristics** | 1 | 1.00 | 1.00 | 1.00 | 1.00 |

**(d) Claude-3-opus (Test Set)**

| Trial | Variable Category | Responses* | Accuracy | Precision | Recall | F1 score |
| --- | --- | --- | --- | --- | --- | --- |
| *GETUG 2015*  *(Gravis et al.)* | **Overall Results** | 1 | 1.00 | 1.00 | 1.00 | 1.00 |
|  | **Population Characteristics** | 1 | 1.00 | 1.00 | 1.00 | 1.00 |
| *GETUG 2013*  *(Gravis et al.)* | **Overall Results** | 1 | 0.00 | 0.00 | - | - |
|  | **Population Characteristics** | 1 | 0.00 | 0.00 | - | - |
| *STAMPEDE 2019*  *(Attard et al.)* | **Overall Results** | 4 | 0.25 | 0.25 | 1.00 | 0.40 |
|  | **Population Characteristics** | 1 | 1.00 | 1.00 | 1.00 | 1.00 |
|  | **Meta-analysis Outcomes** | 3 | 0.00 | 0.00 | 0.00 |  |
| *STAMPEDE 2022*  *(James et al.)* | **Overall Results** | 2 | 0.00 | - | 0.00 | - |
|  | **Population Characteristics** | 2 | 0.00 | - | 0.00 | - |
| *STAMPEDE 2016*  *(James et al.)* | **Overall Results** | 5 | 0.20 | 0.20 | 1.00 | 0.33 |
|  | **Population Characteristics** | 5 | 0.20 | 0.20 | 1.00 | 0.33 |
| *STAMPEDE 2017*  *(James et al.)* | **Overall Results** | 2 | 1.00 | 1.00 | 1.00 | 1.00 |
|  | **Trial Characteristics** | 1 | 1.00 | 1.00 | 1.00 | 1.00 |
|  | **Population Characteristics** | 1 | 1.00 | 1.00 | 1.00 | 1.00 |
| *STAMPEDE 2018*  *(Sydes et al.)* | **Overall Results** | 2 | 0.50 | 1.00 | 0.50 | 0.67 |
|  | **Population Characteristics** | 2 | 0.50 | 1.00 | 0.50 | 0.67 |
| *CHAARTED 2018*  *(Kyriakopoulos*  *et al.)* | **Overall Results** | 1 | 1.00 | 1.00 | 1.00 | 1.00 |
|  | **Population Characteristics** | 1 | 1.00 | 1.00 | 1.00 | 1.00 |
| *SWOG 2022*  *(Agarwal et al.)* | **Overall Results** | 2 | 0.00 | 0.00 | - | - |
|  | **Population Characteristics** | 2 | 0.00 | 0.00 | - | - |
| *PEACE 1 2022*  *(Fizazi et al.)* | **Overall Results** | 8 | 1.00 | 1.00 | 1.00 | 1.00 |
|  | **Trial Characteristics** | 1 | 1.00 | 1.00 | 1.00 | 1.00 |
|  | **Population Characteristics** | 7 | 1.00 | 1.00 | 1.00 | 1.00 |
| *ENZAMET 2019*  *(Davis et al.)* | **Overall Results** | 1 | 0.00 | 0.00 | - | - |
|  | **Meta-analysis Outcomes** | 1 | 0.00 | 0.00 | - | - |
| *TITAN 2021*  *(Chi et al.)* | **Overall Results** | 4 | 1.00 | 1.00 | 1.00 | 1.00 |
|  | **Population Characteristics** | 3 | 1.00 | 1.00 | 1.00 | 1.00 |
|  | **Meta-analysis Outcomes** | 1 | 1.00 | 1.00 | 1.00 | 1.00 |
| *TITAN 2019*  *(Chi et al.)* | **Overall Results** | 7 | 1.00 | 1.00 | 1.00 | 1.00 |
|  | **Population Characteristics** | 4 | 1.00 | 1.00 | 1.00 | 1.00 |
|  | **Meta-analysis Outcomes** | 3 | 1.00 | 1.00 | 1.00 | 1.00 |
| *ARCHES 2022*  *(Armstrong et al.)* | **Overall Results** | 2 | 0.00 | 0.00 | 0.00 | 0.00 |
|  | **Trial Characteristics** | 1 | 0.00 | - | 0.00 | - |
|  | **Population Characteristics** | 1 | 0.00 | - | 0.00 | - |
| *ARASENS 2023*  *(Hussain et al.)* | **Overall Results** | 2 | 0.00 | 0.00 | - | - |
|  | **Meta-analysis Outcomes** | 2 | 0.00 | 0.00 | - | - |
| *ARASENS 2022*  *(Smith et al.)* | **Overall Results** | 5 | 0.00 | 0.00 | 0.00 | - |
|  | **Population Characteristics** | 5 | 0.00 | 0.00 | 0.00 | - |

*Number of discordant responses generated for each variable category by publications

Mean values for accuracy, precision, recall, and F1 score of (a) GPT-4-turbo in prompt development set (b) GPT-4-turbo in test set (c) Claude-3-opus in prompt development set, and (d) Claude-3-opus in test set for the subset of responses where GPT-4-turbo and Claude-3-opus generated discordant responses by publications

### **Supplementary Table 17: Mean evaluation metrics for a subset of responses with concordance after cross-critique by publications**

**(a) Prompt Development Set**

| Trial | Variable Category | Responses* | Accuracy | Precision | Recall | F1 score |
| --- | --- | --- | --- | --- | --- | --- |
| *STAMPEDE 2018*  *(Clarke et al.)* | **Overall Results** | 1 | 1.00 | 1.00 | 1.00 | 1.00 |
|  | **Meta-analysis Outcomes** | 1 | 1.00 | 1.00 | 1.00 | 1.00 |
| *LATITUDE 2017*  *(Fizazi et al.)* | **Overall Results** | 1 | 1.00 | 1.00 | 1.00 | 1.00 |
|  | **Population Characteristics** | 1 | 1.00 | 1.00 | 1.00 | 1.00 |
| *LATITUDE 2019*  *(Fizazi et al.)* | **Overall Results** | 1 | 1.00 | 1.00 | 1.00 | 1.00 |
|  | **Population Characteristics** | 1 | 1.00 | 1.00 | 1.00 | 1.00 |

**(b) Test Set**

| Trial | Variable Category | Responses* | Accuracy | Precision | Recall | F1 score |
| --- | --- | --- | --- | --- | --- | --- |
| *GETUG 2015*  *(Gravis et al.)* | **Overall Results** | 1 | 1.00 | 1.00 | 1.00 | 1.00 |
|  | **Population Characteristics** | 1 | 1.00 | 1.00 | 1.00 | 1.00 |
| *GETUG 2013*  *(Gravis et al.)* | **Overall Results** | 1 | 1.00 | 1.00 | 1.00 | 1.00 |
|  | **Population Characteristics** | 1 | 1.00 | 1.00 | 1.00 | 1.00 |
| *STAMPEDE 2019*  *(Hoyle et al.)* | **Overall Results** | 2 | 0.50 | 0.50 | 1.00 | 0.67 |
|  | **Population Characteristics** | 1 | 1.00 | 1.00 | 1.00 | 1.00 |
|  | **Meta-analysis Outcomes** | 1 | 0.00 | 0.00 | - | - |
| *STAMPEDE 2022*  *(James et al.)* | **Overall Results** | 2 | 0.00 | 0.00 | 0.00 | - |
|  | **Population Characteristics** | 2 | 0.00 | 0.00 | 0.00 | - |
| *STAMPEDE 2016*  *(James et al.)* | **Overall Results** | 1 | 1.00 | 1.00 | 1.00 | 1.00 |
|  | **Population Characteristics** | 1 | 1.00 | 1.00 | 1.00 | 1.00 |
| *SWOG 2022*  *(Agarwal et al.)* | **Overall Results** | 1 | 1.00 | 0.00 | 0.00 | 0.00 |
|  | **Population Characteristics** | 1 | 1.00 | 0.00 | 0.00 | 0.00 |
| *PEACE 1 2022*  *(Fizazi et al.)* | **Overall Results** | 6 | 1.00 | 1.00 | 1.00 | 1.00 |
|  | **Trial Characteristics** | 1 | 1.00 | 1.00 | 1.00 | 1.00 |
|  | **Population Characteristics** | 5 | 1.00 | 1.00 | 1.00 | 1.00 |
| *TITAN 2021*  *(Chi et al.)* | **Overall Results** | 3 | 0.67 | 1.00 | 0.67 | 0.80 |
|  | **Trial Characteristics** | 2 | 0.50 | 1.00 | 0.50 | 0.67 |
|  | **Meta-analysis Outcomes** | 1 | 1.00 | 1.00 | 1.00 | 1.00 |
| *TITAN 2019*  *(Chi et al.)* | **Overall Results** | 2 | 0.50 | 1.00 | 0.50 | 0.67 |
|  | **Population Characteristics** | 2 | 0.50 | 1.00 | 0.50 | 0.67 |
| *ARCHES 2022*  *(Armstrong*  *et al.)* | **Overall Results** | 2 | 0.50 | 1.00 | 0.50 | 0.67 |
|  | **Trial Characteristics** | 1 | 1.00 | 1.00 | 1.00 | 1.00 |
|  | **Population Characteristics** | 1 | 0.00 | - | 0.00 | - |
| *ARASENS 2023*  *(Hussain et al.)* | **Overall Results** | 1 | 1.00 | 0.00 | 0.00 | 0.00 |
|  | **Meta-analysis Outcomes** | 1 | 1.00 | 0.00 | 0.00 | 0.00 |
| *ARASENS 2022*  *(Smith et al.)* | **Overall Results** | 3 | 1.00 | 1.00 | 1.00 | 1.00 |
|  | **Population Characteristics** | 3 | 1.00 | 1.00 | 1.00 | 1.00 |

*Number of concordant responses generated for each variable category after cross-critique by publications

Mean values for accuracy, precision, recall, and F1 score of (a) prompt development and (b) test sets for the subset of responses where GPT-4-turbo and Claude-3-opus generated concordant responses after cross-critique by publications

### **Supplementary Table 18: Mean evaluation metrics for a subset of responses with discordance after cross-critique by publications**

**(a) GPT-4-turbo (Prompt Development Set)**

| Trial | Variable Category | Responses* | Accuracy | Precision | Recall | F1 score |
| --- | --- | --- | --- | --- | --- | --- |
| *ENZAMET 2023*  *(Sweeney et al.)* | **Overall Results** | 2 | 0.00 | 0.00 | 0.00 | - |
|  | **Meta-analysis Outcomes** | 2 | 0.00 | 0.00 | 0.00 | - |

**(b) GPT-4-turbo (Test Set)**

| Trial | Variable Category | Responses* | Accuracy | Precision | Recall | F1 score |
| --- | --- | --- | --- | --- | --- | --- |
| *STAMPEDE 2019*  *(Hoyle et al.)* | **Overall Results** | 2 | 0.00 | 0.00 | - | - |
|  | **Meta-analysis Outcomes** | 2 | 0.00 | 0.00 | - | - |
| *STAMPEDE 2016*  *(James et al.)* | **Overall Results** | 4 | 0.25 | 0.25 | 1.00 | 0.40 |
|  | **Population Characteristics** | 4 | 0.25 | 0.25 | 1.00 | 0.40 |
| *STAMPEDE 2017*  *(James et al.)* | **Overall Results** | 2 | 0.00 | 0.00 | - | - |
|  | **Trial Characteristics** | 1 | 0.00 | 0.00 | - | - |
|  | **Population Characteristics** | 1 | 0.00 | 0.00 | - | - |
| *STAMPEDE 2018*  *(Sydes et al.)* | **Overall Results** | 2 | 0.00 | 0.00 | 0.00 | - |
|  | **Population Characteristics** | 2 | 0.00 | 0.00 | 0.00 | - |
| *CHAARTED 2018*  *(Kyriakopoulos*  *et al.)* | **Overall Results** | 1 | 1.00 | 1.00 | 1.00 | 1.00 |
|  | **Population Characteristics** | 1 | 1.00 | 1.00 | 1.00 | 1.00 |
| *SWOG 2022*  *(Agarwal et al.)* | **Overall Results** | 1 | 0.00 | 0.00 | - | - |
|  | **Population Characteristics** | 1 | 0.00 | 0.00 | - | - |
| *PEACE 1 2022*  *(Fizazi et al.)* | **Overall Results** | 2 | 0.50 | 1.00 | 0.50 | 0.67 |
|  | **Population Characteristics** | 2 | 0.50 | 1.00 | 0.50 | 0.67 |
| *ENZAMET 2019*  *(Davis et al.)* | **Overall Results** | 1 | 0.00 | 0.00 | - | - |
|  | **Meta-analysis Outcomes** | 1 | 0.00 | 0.00 | - | - |
| *TITAN 2021*  *(Chi et al.)* | **Overall Results** | 1 | 0.00 | - | 0.00 | - |
|  | **Population Characteristics** | 1 | 0.00 | - | 0.00 | - |
| *TITAN 2019*  *(Chi et al.)* | **Overall Results** | 5 | 0.40 | 0.50 | 0.67 | 0.57 |
|  | **Population Characteristics** | 2 | 0.50 | 1.00 | 0.50 | 0.67 |
|  | **Meta-analysis Outcomes** | 3 | 0.33 | 0.33 | 1.00 | 0.50 |
| *ARASENS 2023*  *(Hussain et al.)* | **Overall Results** | 1 | 0.00 | 0.00 | - | - |
|  | **Meta-analysis Outcomes** | 1 | 0.00 | 0.00 | - | - |
| *ARASENS 2022*  *(Smith et al.)* | **Overall Results** | 2 | 0.00 | - | 0.00 | - |
|  | **Population Characteristics** | 2 | 0.00 | - | 0.00 | - |

**(c) Claude-3-opus (Prompt Development Set)**

| Trial | Variable Category | Responses* | Accuracy | Precision | Recall | F1 score |
| --- | --- | --- | --- | --- | --- | --- |
| *ENZAMET 2023*  *(Sweeney et al.)* | **Overall Results** | 2 | 0.00 | 0.00 | - | - |
|  | **Meta-analysis Outcomes** | 2 | 0.00 | 0.00 | - | - |

**(d) Claude-3-opus (Test Set)**

| Trial | Variable Category | Responses* | Accuracy | Precision | Recall | F1 score |
| --- | --- | --- | --- | --- | --- | --- |
| *STAMPEDE 2019*  *(Hoyle et al.)* | **Overall Results** | 2 | 0.00 | 0.00 | 0.00 | - |
|  | **Meta-analysis Outcomes** | 2 | 0.00 | 0.00 | 0.00 | - |
| *STAMPEDE 2016*  *(James et al.)* | **Overall Results** | 4 | 0.25 | 0.50 | 0.33 | 0.40 |
|  | **Population Characteristics** | 4 | 0.25 | 0.50 | 0.33 | 0.40 |
| *STAMPEDE 2017*  *(James et al.)* | **Overall Results** | 2 | 1.00 | 1.00 | 1.00 | 1.00 |
|  | **Trial Characteristics** | 1 | 1.00 | 1.00 | 1.00 | 1.00 |
|  | **Population Characteristics** | 1 | 1.00 | 1.00 | 1.00 | 1.00 |
| *STAMPEDE 2018*  *(Sydes et al.)* | **Overall Results** | 2 | 0.50 | 0.50 | 1.00 | 0.67 |
|  | **Population Characteristics** | 2 | 0.50 | 0.50 | 1.00 | 0.67 |
| *CHAARTED 2018*  *(Kyriakopoulos*  *et al.)* | **Overall Results** | 1 | 0.00 | 0.00 | - | - |
|  | **Population Characteristics** | 1 | 0.00 | 0.00 | - | - |
| *SWOG 2022*  *(Agarwal et al.)* | **Overall Results** | 1 | 1.00 | 1.00 | 1.00 | 1.00 |
|  | **Population Characteristics** | 1 | 1.00 | 1.00 | 1.00 | 1.00 |
| *PEACE 1 2022*  *(Fizazi et al.)* | **Overall Results** | 2 | 0.50 | 0.50 | 1.00 | 0.67 |
|  | **Population Characteristics** | 2 | 0.50 | 0.50 | 1.00 | 0.67 |
| *ENZAMET 2019*  *(Davis et al.)* | **Overall Results** | 1 | 0.00 | 0.00 | - | - |
|  | **Meta-analysis Outcomes** | 1 | 0.00 | 0.00 | - | - |
| *TITAN 2021*  *(Chi et al.)* | **Overall Results** | 1 | 1.00 | 1.00 | 1.00 | 1.00 |
|  | **Population Characteristics** | 1 | 1.00 | 1.00 | 1.00 | 1.00 |
| *TITAN 2019*  *(Chi et al.)* | **Overall Results** | 5 | 0.80 | 1.00 | 0.80 | 0.89 |
|  | **Population Characteristics** | 2 | 0.50 | 1.00 | 0.50 | 0.67 |
|  | **Meta-analysis Outcomes** | 3 | 1.00 | 1.00 | 1.00 | 1.00 |
| *ARASENS 2023*  *(Hussain et al.)* | **Overall Results** | 1 | 0.00 | 0.00 | - | - |
|  | **Meta-analysis Outcomes** | 1 | 0.00 | 0.00 | - | - |
| *ARASENS 2022*  *(Smith et al.)* | **Overall Results** | 2 | 0.00 | 0.00 | - | - |
|  | **Population Characteristics** | 2 | 0.00 | 0.00 | - | - |

*Number of discordant responses generated for each variable category after cross-critique by publications

Mean values for accuracy, precision, recall, and F1 score of (a) GPT-4-turbo in prompt development set (b) GPT-4-turbo in test set (c) Claude-3-opus in prompt development set, and (d) Claude-3-opus in test set for the subset of responses where GPT-4-turbo and Claude-3-opus generated discordant responses after cross-critique by publications

### **Supplementary Table 19: Mean percentage hallucinations with 95% confidence intervals for responses across all publications**

**(a) GPT-4-turbo in a zero-shot setting**

| Variable Category | Development | Test |
| --- | --- | --- |
| Overall Results | 0.00 (-) | 2.29 (0.61-3.98) |
| Trial Characteristics | 0.00 (-) | 0.00 (-) |
| Population Characteristics | 0.00 (-) | 3.18 (0.17-6.18) |
| Meta-analysis Outcomes | 0.00 (-) | 1.59 (0.00-3.85) |

**(b) Claude-3-opus in a zero-shot setting**

| Variable Category | Development | Test |
| --- | --- | --- |
| Overall Results | 2.60 (0.00-6.08) | 2.76 (0.27-5.26) |
| Trial Characteristics | 0.00 (-) | 0.00 (-) |
| Population Characteristics | 0.00 (-) | 3.71 (0.00-8.26) |
| Meta-analysis Outcomes | 5.40 (0.00-7.06) | 4.06 (0.00-10.99) |

Mean values with corresponding 95% confidence intervals for percentage hallucinations of (a) GPT-4-turbo and (b) Claude 3-opus for all the responses of all publications in a zero-shot setting

### **Supplementary Table 20: Mean percentage hallucinations with 95% confidence intervals for a subset of responses with concordance**

| Variable Category | Development | Test |
| --- | --- | --- |
| Overall Results | 0.00 (-) | 0.24 (0.00-0.70) |
| Trial Characteristics | 0.00 (-) | 0.00 (-) |
| Population Characteristics | 0.00 (-) | 0.00 (-) |
| Meta-analysis Outcomes | 0.00 (-) | 0.53 (0.00-1.03) |

Mean values with corresponding 95% confidence intervals for percentage hallucinations for the subset of responses where GPT-4-turbo and Claude-3-opus generated concordant responses

### **Supplementary Table 21: Mean percentage hallucinations with 95% confidence intervals for a subset of responses with discordance**

**(a) GPT-4-turbo in a zero-shot setting**

| Variable Category | Development | Test |
| --- | --- | --- |
| Overall Results | 0.00 (-) | 26.93 (6.38-47.48) |
| Trial Characteristics | 0.00 (-) | 0.00 (-) |
| Population Characteristics | 0.00 (-) | 30.77 (7.13-54.41) |
| Meta-analysis Outcomes | 0.00 (-) | 20.00 (0.00-59.20) |

**(a) Claude-3-opus in a zero-shot setting**

| Variable Category | Development | Test |
| --- | --- | --- |
| Overall Results | 50.00 (0.00-100.00) | 41.00 (17.41-64.59) |
| Trial Characteristics | 0.00 (-) | 0.00 (-) |
| Population Characteristics | 0.00 (-) | 26.20 (3.36-48.95) |
| Meta-analysis Outcomes | 100.00 (-) | 60.00 (11.99-100.00) |

Mean values with corresponding 95% confidence intervals for percentage hallucinations of (a) GPT-4-turbo and (b) Claude-3-opus for the subset of responses where GPT-4-turbo and Claude-3-opus generated discordant responses

### **Supplementary Table 22: Mean percentage hallucinations with 95% confidence intervals for a subset of responses with concordance after cross-critique**

| Variable Category | Development | Test |
| --- | --- | --- |
| Overall Results | 0.00 (-) | 8.30 (0.00-19.35) |
| Trial Characteristics | 0.00 (-) | 0.00 (-) |
| Population Characteristics | 0.00 (-) | 4.50 (0.00-13.45) |
| Meta-analysis Outcomes | 0.00 (-) | 33.33 (0.00-98.67) |

Mean values with corresponding 95% confidence intervals for percentage hallucinations for the subset of responses where GPT-4-turbo and Claude-3-opus generated concordant responses after cross-critique

### **Supplementary Table 23: Mean percentage hallucinations with 95% confidence intervals for a subset of responses with discordance after cross-critique**

**(a) GPT-4-turbo after cross-critique**

| Variable Category | Development | Test |
| --- | --- | --- |
| Overall Results | 50.00 (-) | 56.25 (30.57-81.93) |
| Trial Characteristics | - | - |
| Population Characteristics | - | 36.11 (6.54-65.68) |
| Meta-analysis Outcomes | 50.00 (-) | 100.00 (-) |

**(b) Claude-3-opus after cross-critique**

| Variable Category | Development | Test |
| --- | --- | --- |
| Overall Results | 100.00 (-) | 47.92 (20.64-75.20) |
| Trial Characteristics | - | - |
| Population Characteristics | - | 41.67 (11.11-72.22) |
| Meta-analysis Outcomes | 100.00 (-) | 50.00 (0.00-100.00) |

Mean values with corresponding 95% confidence intervals for percentage hallucinations of (a) GPT-turbo and (b) Claude-3-opus for the subset of responses where GPT-4-turbo and Claude-3-opus generated discordant responses after cross-critique

### **Supplementary Table 24: Sensitivity analysis showing mean percentage hallucinations with 95% confidence intervals for responses across all publications of clinical trials included exclusively in the test set**

**(a) GPT-4-turbo in a zero-shot setting**

| Variable Category | Test |
| --- | --- |
| Overall Results | 2.00 (0.00-4.59) |
| Trial Characteristics | 0.00 (-) |
| Population Characteristics | 2.00 (0.00-5.92) |
| Meta-analysis Outcomes | 2.00 (0.00-5.92) |

**(b) Claude-3-opus in a zero-shot setting**

| Variable Category | Test |
| --- | --- |
| Overall Results | 1.44 (0.00-3.49) |
| Trial Characteristics | 0.00 (-) |
| Population Characteristics | 3.00 (0.00-7.16) |
| Meta-analysis Outcomes | 0.00 (-) |

Sensitivity analysis showing mean values with corresponding 95% confidence intervals for percentage hallucinations of (a) GPT-4-turbo and (b) Claude-3-opus for the responses of all publications of clinical trials included exclusively in the test set in a zero-shot setting

### **Supplementary Table 25: Sensitivity analysis showing mean percentage hallucinations with 95% confidence intervals for responses with concordance across publications of clinical trials included exclusively in the test set**

| Variable Category | Test |
| --- | --- |
| Overall Results | 0.00 (-) |
| Trial Characteristics | 0.00 (-) |
| Population Characteristics | 0.00 (-) |
| Meta-analysis Outcomes | 0.00 (-) |

Sensitivity analysis showing mean values with corresponding 95% confidence intervals for percentage hallucinations for the subset of responses where GPT-4-turbo and Claude-3-opus generated concordant responses across publications of clinical trials included exclusively in the test set

### **Supplementary Table 26: Sensitivity analysis showing mean percentage hallucinations with 95% confidence intervals for responses with discordance across publications of clinical trials included exclusively in the test set**

**(a) GPT-4-turbo in a zero-shot setting**

| Variable Category | Test |
| --- | --- |
| Overall Results | 14.33 (0.00-36.24) |
| Trial Characteristics | 0.00 (-) |
| Population Characteristics | 18.75 (0.00-44.53) |
| Meta-analysis Outcomes | 0.00 (-) |

**(b) Claude-3-opus in a zero-shot setting**

| Variable Category | Test |
| --- | --- |
| Overall Results | 50.00 (6.17-98.83) |
| Trial Characteristics | 0.00 (-) |
| Population Characteristics | 40.00 (0.00-88.01) |
| Meta-analysis Outcomes | 50.00 (0.00-98.00) |

Sensitivity analysis showing mean values with corresponding 95% confidence intervals for percentage hallucinations of (a) GPT-4-turbo and (b) Claude 3-opus for a subset of responses where GPT-4-turbo and Claude-3-opus generated discordant responses across publications of clinical trials included exclusively in the test set

### **Supplementary Table 27: Sensitivity analysis showing mean percentage hallucinations with 95% confidence intervals for responses with concordance after cross-critique across publications of clinical trials included exclusively in the test set**

| Variable Category | Test |
| --- | --- |
| Overall Results | 0.00 (-) |
| Trial Characteristics | 0.00 (-) |
| Population Characteristics | 0.00 (-) |
| Meta-analysis Outcomes | 0.00 (-) |

Mean values with corresponding 95% confidence intervals for percentage hallucinations for the subset of responses where GPT-4-turbo and Claude-3-opus generated concordant responses after cross-critique across publications of clinical trials included exclusively in the test set

### **Supplementary Table 28: Sensitivity analysis showing mean percentage hallucinations with 95% confidence intervals for responses with discordance after cross-critique across publications of clinical trials included exclusively in the test set**

**(a) GPT-4-turbo after cross-critique**

| Variable Category | Test |
| --- | --- |
| Overall Results | 14.67 (2.33-81.00) |
| Trial Characteristics | - |
| Population Characteristics | 20.00 (0.00-59.20) |
| Meta-analysis Outcomes | 100.00 (-) |

**(b) Claude-3-opus after cross-critique**

| Variable Category | Test |
| --- | --- |
| Overall Results | 50.00 (6.17-93.83) |
| Trial Characteristics | - |
| Population Characteristics | 40.00 (0.00-88.01) |
| Meta-analysis Outcomes | 50.00 (0.00-100.00) |

Mean values with corresponding 95% confidence intervals for percentage hallucinations of (a) GPT-turbo and (b) Claude-3-opus for the subset of responses where GPT-4-turbo and Claude-3-opus generated discordant responses after cross-critique across publications of clinical trials included exclusively in the test set

### **Supplementary Figure 1: Prompt engineering workflow for data extraction**

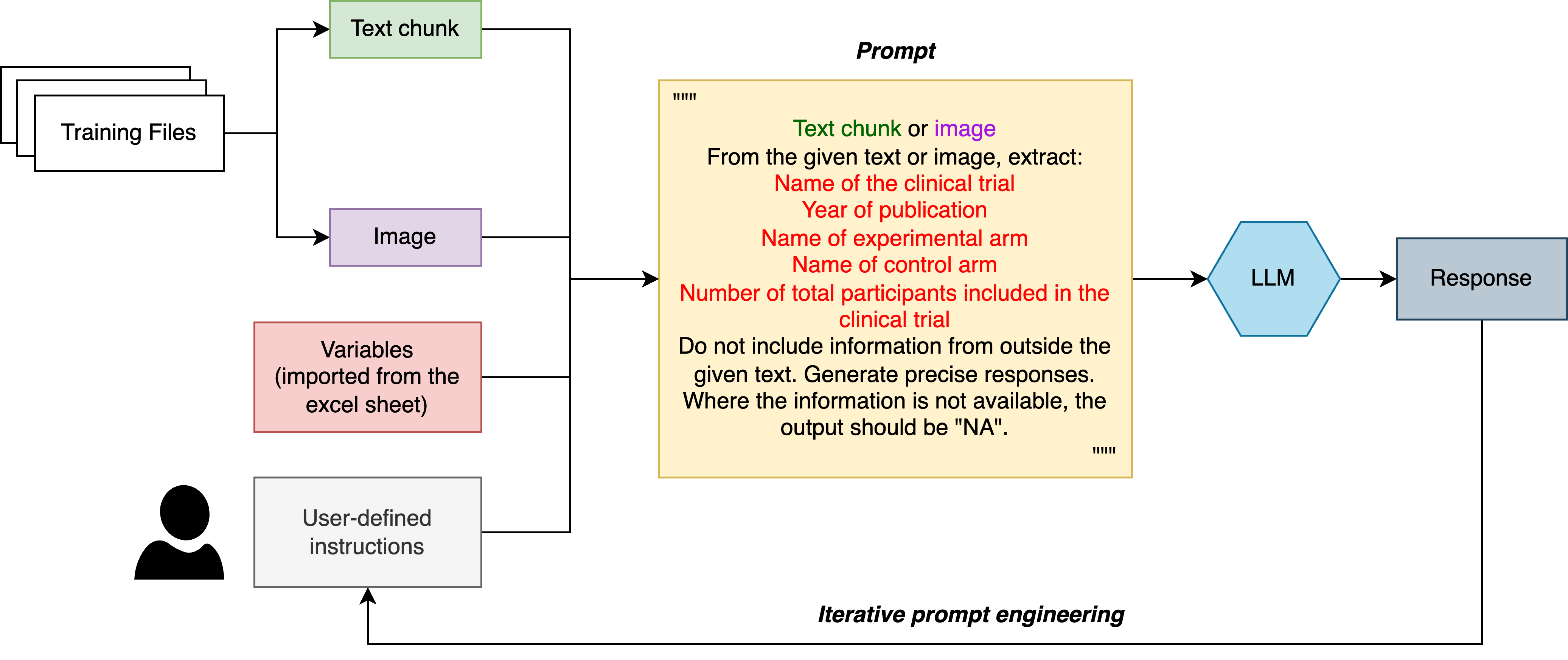

Abbreviations: LLM: large language model

The figure shows the schema for structuring and engineering the prompts used for data extraction. It explains the process with an example where the prompt begins with the extracted chunk of text or image. Text chunks and images are programmatically processed from the PDF files of clinical trial reports. This is followed by the names of variables for which data extraction is required. The proposed approach allows the flexibility to add, remove, and edit existing variables in an external spreadsheet, which can be automatically incorporated within the prompt. This is followed by user-defined instructions to generate precise responses, generating “NA” where the responses are not available. Based on the LLM-generated responses, the prompts can be iteratively modified to extract the variables required for a systematic review

### **Supplementary Figure 2: Breakdown of the generated responses**

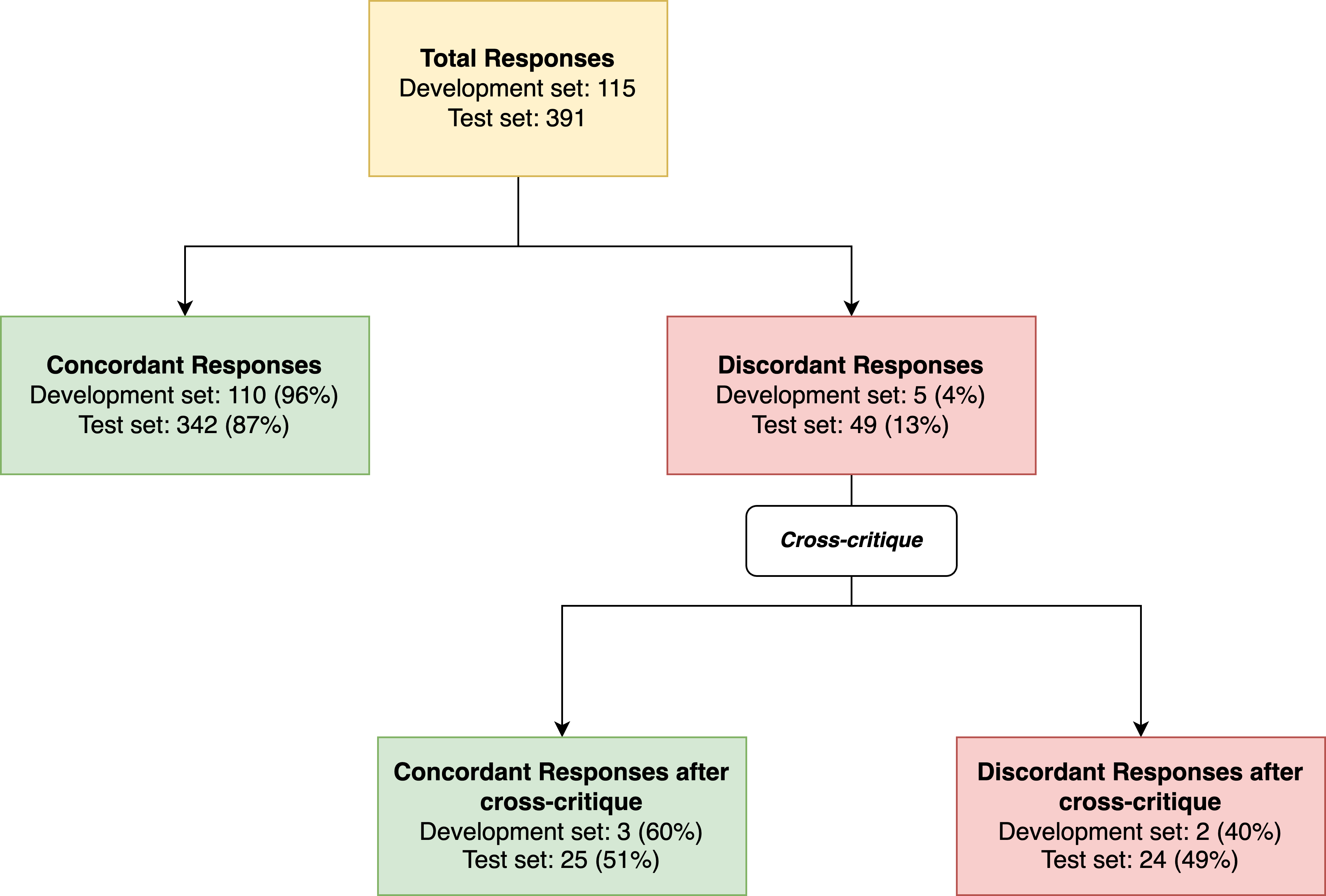

The figure shows the breakdown of responses generated by GPT-4-turbo and Claude-3-opus, showing the number of concordant and discordant responses before and after cross-critique. The responses where GPT-4-turbo and Claude-3-opus generated similar responses were said to exhibit concordance. The breakdown of responses shows the majority of responses (96% in the prompt development set; 87% in the test set) exhibiting concordance. Cross-critique shows the subset of regenerated responses to demonstrate concordance where the responses were initially discordant

### **Supplementary Figure 3: Mean precision of the individual versus the 2-reviewer collaborative LLM approach in the test set responses**

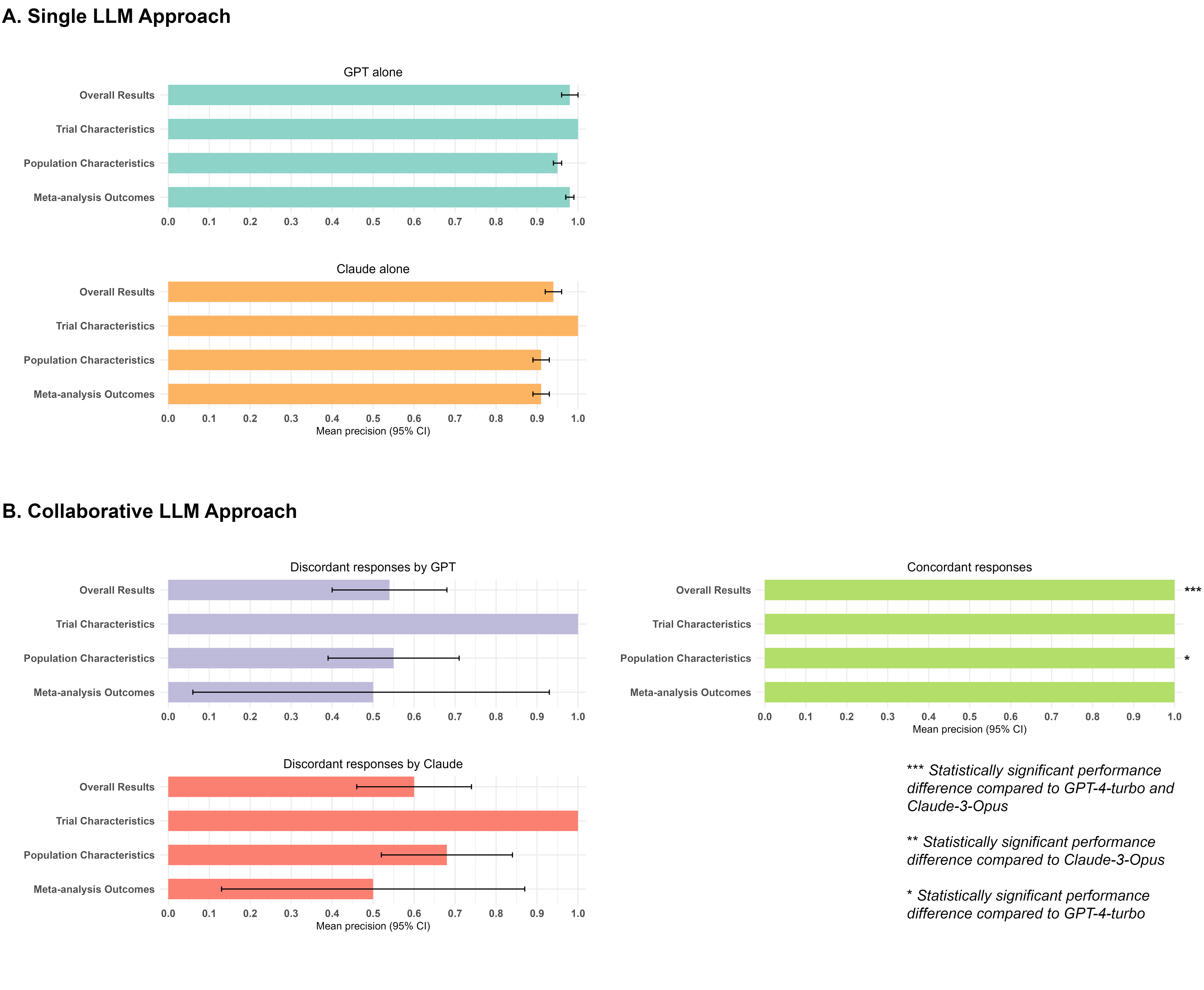

Abbreviations: LLM: Large language model; GPT: Generative pretrained transformer

The figure shows (A). Mean data extraction precision with 95% confidence intervals (CI) of GPT-4-turbo and Claude-3-Opus in the single LLM approach. (B). Mean data extraction precision with 95% CI of the discordant responses by GPT-4-turbo and Claude-3-Opus, and for concordant responses in the collaborative LLM approach. The concordant responses significantly outperform either GPT-4-turbo or Claude-3-Opus alone in overall data extraction precision and outperforms GPT-4-turbo in precision for extracting population characteristics from clinical trial publications

### **Supplementary Figure 4: Mean recall of the individual versus the 2-reviewer collaborative LLM approach in the test set responses**

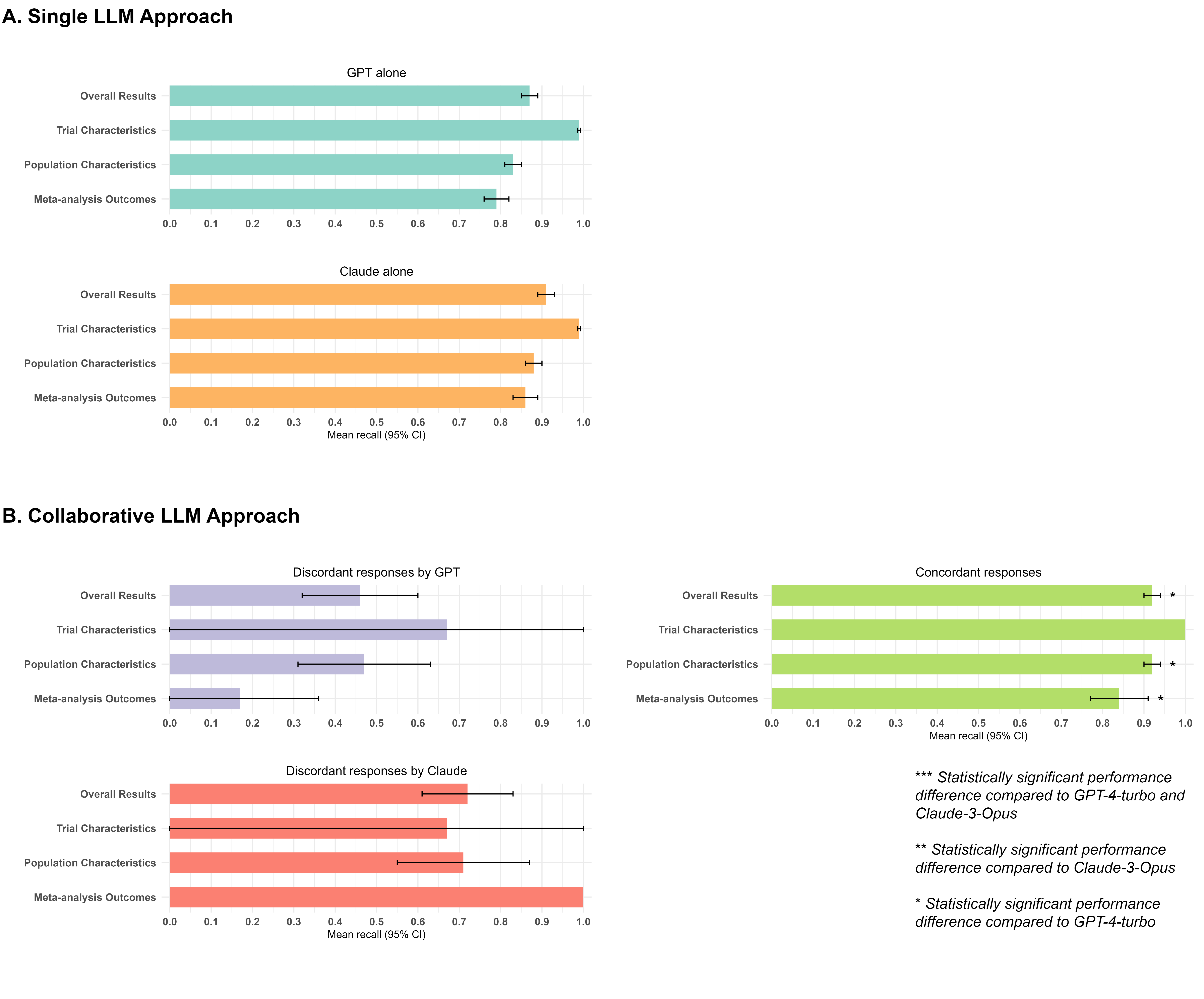

Abbreviations: LLM: Large language model; GPT: Generative pretrained transformer

The figure shows (A). Mean data extraction recall with 95% confidence intervals (CI) of GPT-4-turbo and Claude-3-Opus in the single LLM approach. (B). Mean data extraction recall with 95% CI of the discordant responses by GPT-4-turbo and Claude-3-Opus, and for concordant responses in the collaborative LLM approach. The concordant responses significantly outperform GPT-4-turbo alone in overall data extraction recall and recall for extracting population characteristics and meta-analysis outcomes from clinical trial publications

### **Supplementary Figure 5: Mean F1 scores of the individual versus the 2-reviewer collaborative LLM approach in the test set responses**

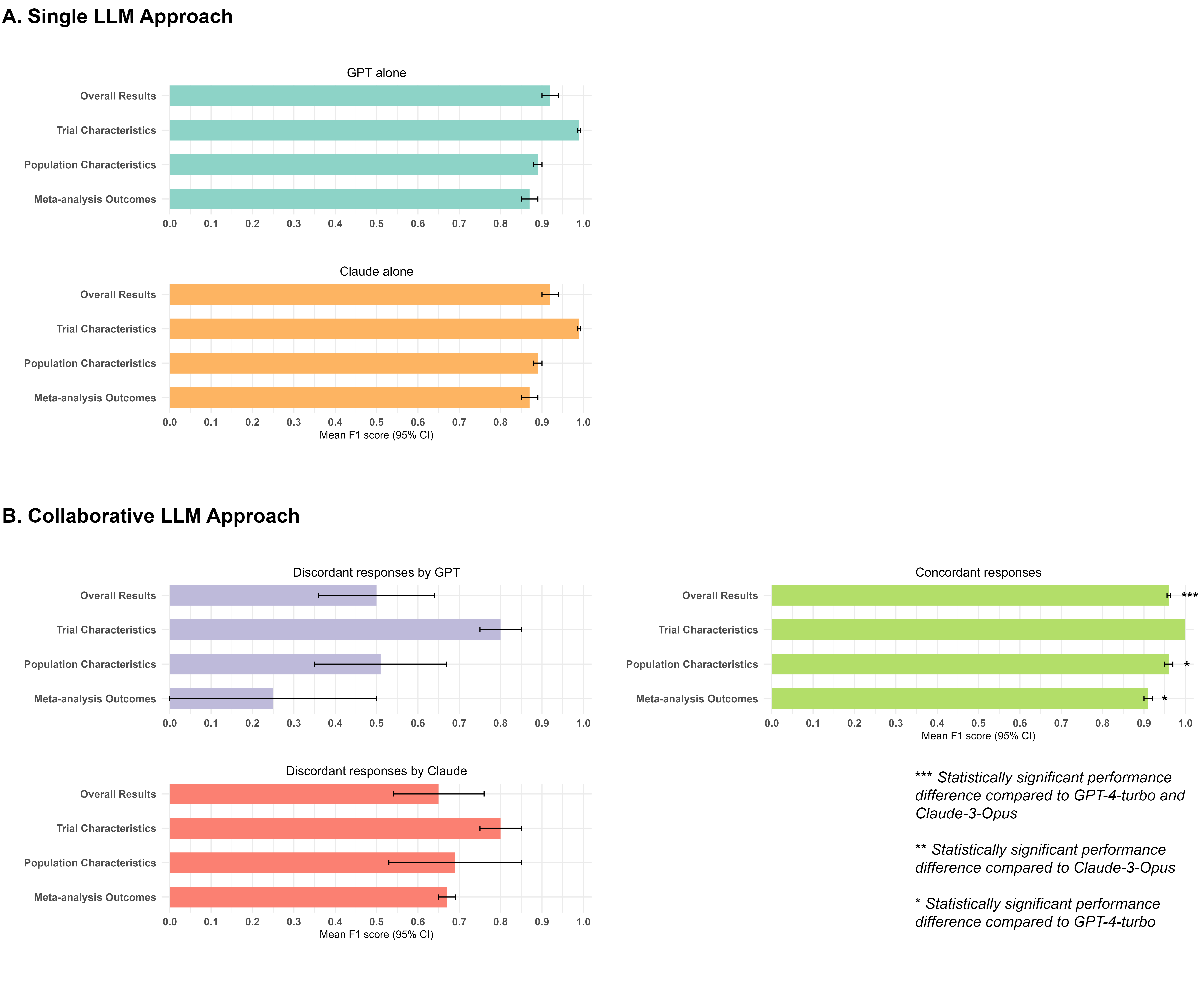

Abbreviations: LLM: Large language model; GPT: Generative pretrained transformer

The figure shows (A). Mean data extraction F1 score with 95% confidence intervals (CI) of GPT-4-turbo and Claude-3-Opus in the single LLM approach. (B). Mean data extraction F1 score with 95% CI of the discordant responses by GPT-4-turbo and Claude-3-Opus, and for concordant responses in the collaborative LLM approach. The concordant responses significantly outperform either GPT-4-turbo or Claude-3-Opus alone in overall data extraction F1 score and outperforms GPT-4-turbo in F1 score for extracting population characteristics and meta-analysis outcomes from clinical trial publications

### **Supplementary Figure 6: Mean precision of the test set discordant responses versus the concordant responses after cross-critique**

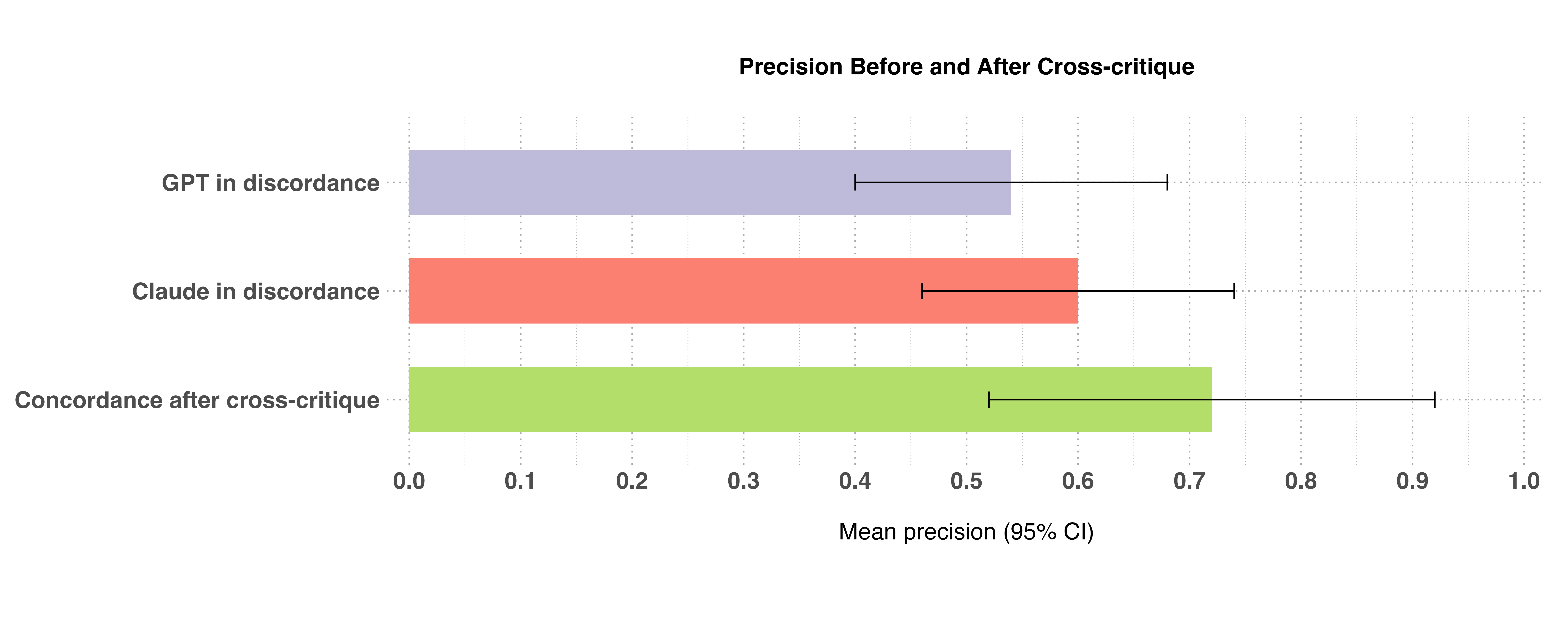

Abbreviations: GPT: Generative pre-trained transformer

The figure shows mean test set precision with 95% confidence intervals of the discordant responses by GPT-4-turbo and Claude-3-Opus, and the concordant responses after the cross-critique. Cross-critique has shown to generate concordant responses with a higher precision than the discordant responses by either GPT-4-turbo or Claude-3-Opus

### **Supplementary Figure 7: Mean recall of the test set discordant responses versus the concordant responses after cross-critique**

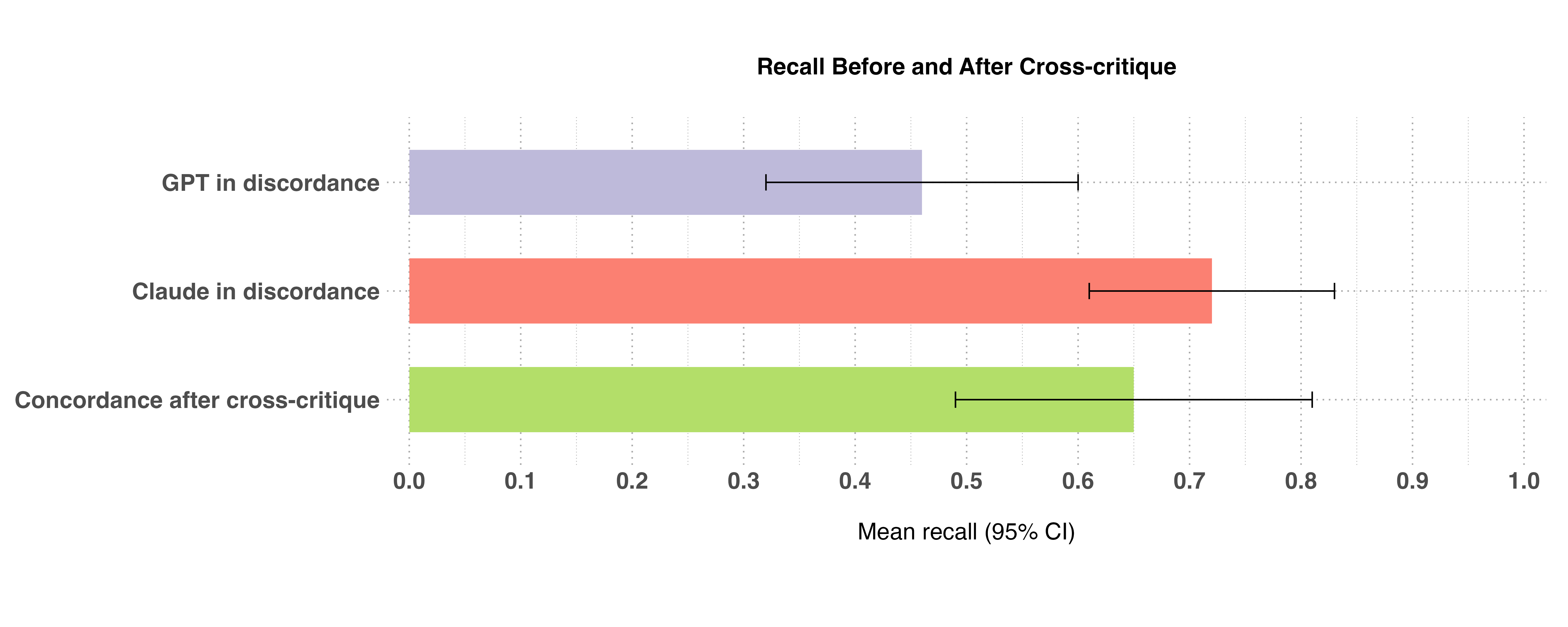

Abbreviations: GPT: Generative pre-trained transformer

The figure shows mean test set recall with 95% confidence intervals of the discordant responses by GPT-4-turbo and Claude-3-Opus, and the concordant responses after the cross-critique. Cross-critique has shown to generate concordant responses with a higher recall than the discordant responses by GPT-4-turbo. Discordant responses by Claude-3-Opus have the highest mean recall.

### **Supplementary Figure 8: Mean F1 scores of the test set discordant responses versus the concordant responses after cross-critique**

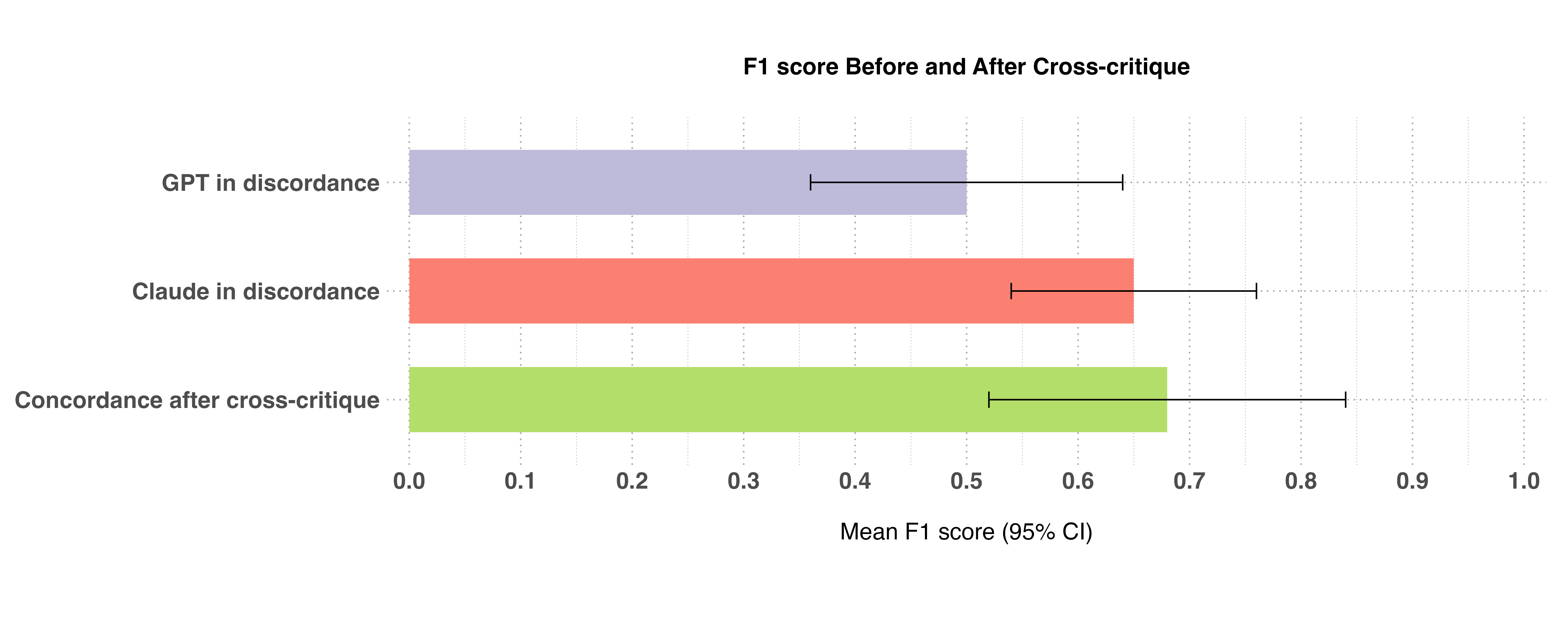

Abbreviations: GPT: Generative pre-trained transformer

The figure shows mean test set F1 score with 95% confidence intervals of the discordant responses by GPT-4-turbo and Claude-3-Opus, and the concordant responses after the cross-critique. Cross-critique has shown to generate concordant responses with a higher F1-score than the discordant responses by either GPT-4-turbo or Claude-3-Opus
